# Hepatic insulin resistance is the basis of bile acid dysmetabolism in metabolic dysfunction-associated steatotic liver disease

**DOI:** 10.1101/2025.11.18.25340381

**Authors:** Sami F. Qadri, Joel T. Haas, Sirkku Jäntti, Kimmo Porthan, Anne Juuti, Anne K. Penttilä, Johanna Arola, Emilia Vartiainen, Eveline Dirinck, Luisa Vonghia, Sven Francque, Matej Orešič, Taru Tukiainen, Tuulia Hyötyläinen, Bart Staels, Hannele Yki-Järvinen

## Abstract

Bile acids (BAs) are liver-synthesized steroids that facilitate lipid digestion and regulate diverse metabolic pathways. Because of their cytotoxicity, excessive hepatic BA accumulation has been implicated in the pathogenesis of metabolic dysfunction-associated steatotic liver disease (MASLD). Although circulating BAs are frequently elevated in MASLD, it is unclear whether similar alterations occur within the liver. Moreover, the strong overlap between MASLD and metabolic syndrome complicates efforts to distinguish BA changes driven by liver disease from those arising due to broader metabolic dysfunction. Here we show in a series of human studies that the BA dysmetabolism in MASLD originates from insulin resistance rather than hepatic steatosis. We found that circulating BA concentrations are twofold higher in patients with MASLD than in healthy controls, despite similar intrahepatic levels. Causal inference using a MASLD genetic risk score indicated that this elevation is not attributable to hepatic steatosis. Instead, circulating BAs, but not intrahepatic BAs, associate with glycemia and hepatic insulin sensitivity. We found reduced expression of the BA uptake transporter NTCP in insulin-resistant individuals, implicating impaired hepatic BA clearance. During hepatic vein catheterization, insulin acutely lowered conjugated BA concentrations in hepatic venous blood, consistent with diminished splanchnic BA spillover. Physiology-based simulations of impaired hepatic BA clearance recapitulated the human phenotype. Taken together, our findings argue against a major role for BAs in MASLD pathogenesis but reveal a previously unrecognized link between circulating BA dynamics and hepatic insulin action.

## Introduction

Metabolic dysfunction-associated steatotic liver disease (MASLD) is the most common chronic liver disorder and can progress to steatohepatitis (MASH) and cirrhosis.^1^ Intrahepatic triglycerides (IHTGs) accumulate in response to chronic overnutrition and physical inactivity, with strong genetic influence.^2^ A small number of common variants account for a third of the population-level variance in IHTG.^3^ These genetic instruments have emerged as powerful tools for dissecting causal relationships between hepatic steatosis its metabolic sequelae.^4–7^

Bile acids (BAs) are amphipathic steroid derivatives that facilitate intestinal lipid absorption and can also act as potent metabolic signaling molecules.^8,9^ The human liver synthesizes the primary BAs chenodeoxycholic acid (CDCA) and cholic acid (CA) from cholesterol, rapidly conjugates them with glycine or taurine, and secretes them into the duodenum (illustrated in **Fig. 1A**).^10^ Gut microbiota subsequently deconjugate and transform primary BAs into many secondary species, predominantly deoxycholic acid (DCA).^11^ Approximately 95% of intestinal BAs are reabsorbed and returned to the liver via the portal vein, where hepatocytes clear them efficiently through the sodium/taurocholate co-transporting polypeptide (NTCP).^12,13^ However, a small fraction escapes hepatic uptake and spills over into the systemic circulation.^10^ Circulating BAs activate the farnesoid X receptor (FXR) and the Takeda G protein-coupled receptor 5 (TGR5) in extrahepatic tissues to regulate glucose, lipid, and energy homeostasis.^8^

**Figure 1.**
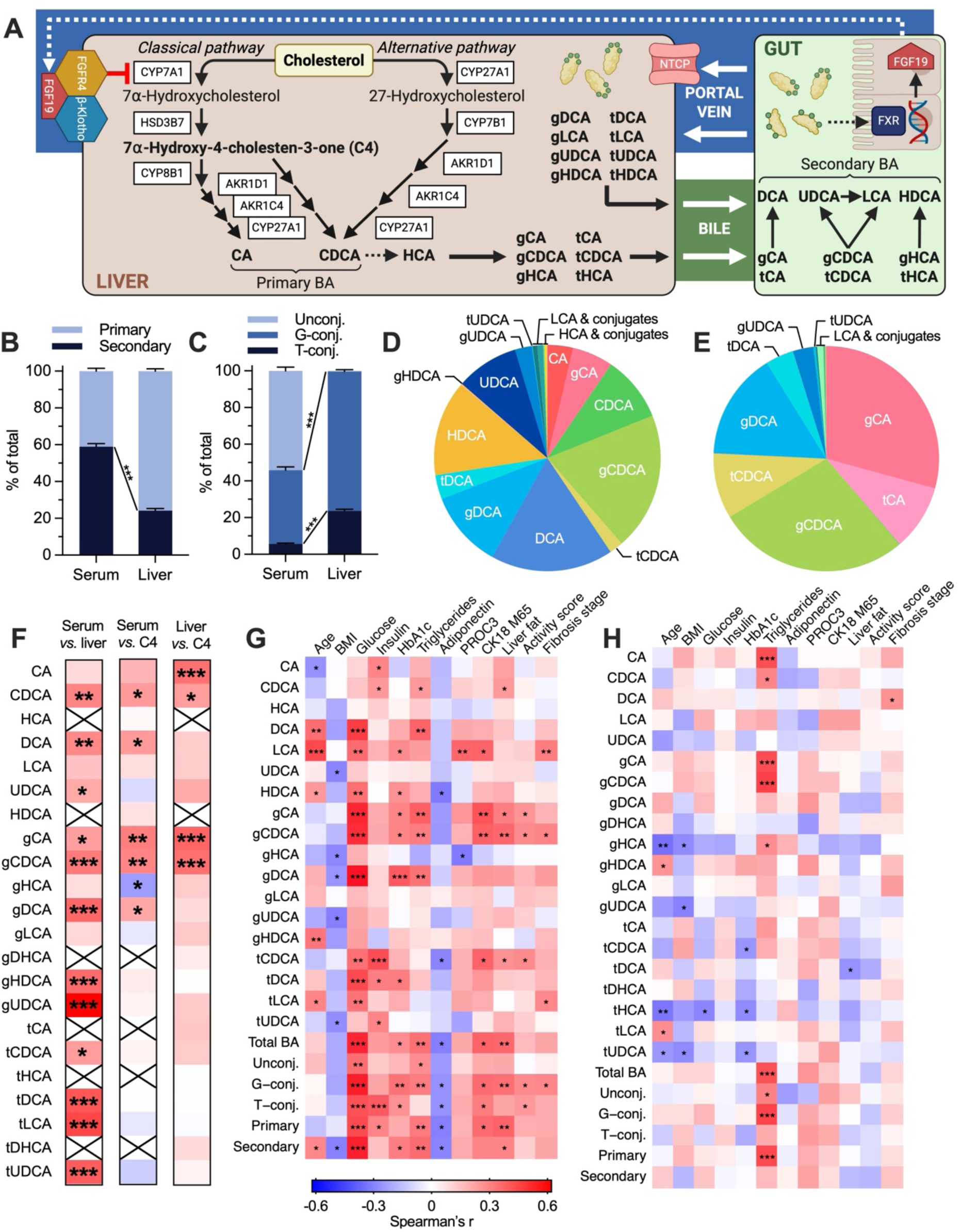
Composition of circulating and hepatic BA pools and their associations with features of MASLD. **(A)** Biosynthesis, enterohepatic circulation, and negative feedback regulation of BA metabolism. Intestinal BA absorption activates FXR in enterocytes, inducing transcription of fibroblast growth factor 19 (FGF19).^37^ Secreted FGF19 reaches the liver via mesenteric blood, where it binds fibroblast growth factor receptor 4 (FGFR4) and its co-receptor β-Klotho, repressing CYP7A1 and thereby inhibiting BA synthesis.^37,38^ (**B**–**C**) Distribution of (**B**) primary *vs.* secondary BAs and (**C**) unconjugated *vs.* conjugated BAs in serum and liver. The Student’s *t*-test was used. Data are mean ± SEM. (**D**–**E**) Relative abundance of individual BAs (**D**) in serum and (**E**) in liver. Unlabeled BAs either comprised <0.1% of the total pool or were undetectable. (**F**) Heatmaps illustrating correlations between serum *vs.* liver BAs (left), serum BAs *vs.* serum C4 (middle), and liver BAs *vs.* serum C4 (right). Crossed cells indicate undetected BAs. (**G**–**H**) Heatmaps illustrating correlations between (**G**) serum and (**H**) liver BAs and key clinical and histological parameters. \**P* < 0.05; \*\**P* < 0.01; \*\*\**P* < 0.001.

Given their detergent-like and cytotoxic properties, dysregulated BA metabolism is implicated in MASLD-related liver injury.^14–17^ Circulating BAs are frequently elevated in MASLD, although reported compositional changes are inconsistent.^18^ Only two small studies have quantified intrahepatic BAs, yielding discordant results.^19,20^ Interpreting these observations is challenging because steatosis is tightly linked to metabolic dysfunction.^21^ Indeed, BA alterations resembling those reported in MASLD also occur in obesity, metabolic syndrome, and type 2 diabetes,^22–28^ making it difficult to distinguish effects of IHTG from those of insulin resistance. Moreover, many studies assessing BAs in MASLD did not evaluate glucose metabolism or insulin sensitivity^15,29,30^, and where this was done, substantial between-group differences were evident.^15,31–33^ BMI likewise differed markedly in most comparisons.^15,29–33^ Emerging evidence suggests that circulating BA changes track more closely with metabolic dysfunction than with features of MASH.^34,35^ Yet, no study has fully disentangled the independent effects of IHTG and insulin resistance on BA metabolism, leaving fundamental questions unresolved.

Here, we conducted a series of human studies to determine whether MASLD-related changes in BA pool size and composition arise from hepatic steatosis or insulin resistance. We confirm a striking increase in circulating BAs in MASLD, without corresponding changes in intrahepatic BAs. Using a genetic risk score (GRS) for IHTG content to infer causality,^4^ we show that this elevation is not attributable to liver steatosis. Instead, circulating BAs correlate strongly with glycemia and hepatic insulin sensitivity. In liver biopsies, we identify reduced NTCP expression in insulin-resistant individuals with MASLD, suggesting impaired hepatic BA uptake. We further show that insulin infusion acutely suppresses conjugated BAs in hepatic venous blood, consistent with a regulatory role of insulin in splanchnic BA spillover. Finally, a physiology-based simulation of human BA metabolism reproduces the *in vivo* findings, supporting impaired hepatic BA clearance as a plausible mechanism underlying systemic BA elevations.

## Results

### Hepatic and circulating BAs are interrelated, but only circulating BAs correlate with features of MASLD

To investigate how MASLD influences hepatic and circulating BA metabolism, we profiled BAs in a deeply phenotyped liver biopsy cohort of 103 overweight or obese individuals (**Table 1**). The participants had a wide range of IHTG content spanning 0% to 80%. MASLD was present in 75% and MASH in 10%. Fibrosis was generally mild: 34% had stage F1 and 5% had stage F2, with no cases of advanced fibrosis (F3–F4).

**Table 1.** Clinical characteristics of the main liver biopsy cohort and subgroups stratified by HOMA-IR and the GRS.

|  | All patients | Low HOMA-IR | High HOMA-IR | Low GRS | High GRS |
| --- | --- | --- | --- | --- | --- |
| Group size, *n* | 103 | 51 | 52 | 50 | 53 |
| Age, years | 48 ± 1 | 49 ± 1 | 47 ± 1 | 46 ± 1 | 50 ± 1* |
| Females, *n* (%) | 69 (67) | 34 (67) | 35 (67) | 33 (66) | 36 (68) |
| BMI, kg/m<sup>2</sup> | 45.3 (41.3–49.1) | 43.6 (40.4–49.3) | 45.4 (43.3–49.1) | 45.2 (40.7–49.2) | 45.4 (41.6–49.0) |
| Waist circumference, cm | 129 (121–140) | 127 (118–137) | 134 (126–141)* | 128 (120–139) | 129 (123–140) |
| Systolic blood pressure, mmHg | 134 (122–145) | 133 (124–147) | 135 (121–141) | 128 (121–144) | 137 (126–145) |
| Diastolic blood pressure, mmHg | 90 (84–97) | 90 (84–95) | 92 (84–97) | 88 (82–95) | 93 (85–98) |
| Plasma glucose, mmol/L | 5.7 (5.2–6.5) | 5.5 (4.8–6.0) | 6.1 (5.6–7.0)*** | 5.7 (5.2–6.3) | 5.8 (5.3–6.7) |
| HbA1c, % | 5.9 (5.6–6.3) | 5.7 (5.6–6.1) | 6.0 (5.7–6.6)* | 5.9 (5.6–6.3) | 6.0 (5.6–6.2) |
| Serum insulin, mIU/L | 12.0 (7.2–17.2) | 7.4 (6.1–10.4) | 17.2 (13.9–21.5)*** | 11.9 (7.5–17.1) | 12.3 (6.9–17.3) |
| Serum C-peptide, nmol/L | 1.09 ± 0.04 | 0.84 ± 0.04 | 1.33 ± 0.05*** | 1.13 ± 0.06 | 1.05 ± 0.05 |
| HOMA-IR | 3.2 (1.8–4.7) | 1.8 (1.3–2.6) | 4.7 (3.7–5.7)*** | 3.2 (1.8–4.8) | 3.1 (1.8–4.6) |
| Matsuda ISI† | 52.0 (36.6–84.1) | 84.1 (63.7–120.6) | 36.6 (28.3–47.1)*** | 51.7 (29.6–82.1) | 52.3 (39.4–84.8) |
| Plasma total cholesterol, mmol/L | 4.1 (3.5–4.7) | 3.9 (3.5–4.7) | 4.2 (3.7–4.7) | 4.2 (3.5–4.7) | 4.0 (3.6–4.7) |
| Plasma HDL cholesterol, mmol/L | 1.11 (0.96–1.29) | 1.11 (0.97–1.37) | 1.04 (0.94–1.23) | 1.11 (0.96–1.32) | 1.11 (0.95–1.27) |
| Plasma LDL cholesterol, mmol/L | 2.3 (1.7–2.9) | 2.3 (1.6–2.8) | 2.4 (1.9–3.1) | 2.3 (1.7–3.0) | 2.4 (1.6–2.9) |
| Plasma triglycerides, mmol/L | 1.26 (1.02–1.73) | 1.14 (0.95–1.69) | 1.31 (1.10–1.92) | 1.29 (0.97–1.88) | 1.22 (1.04–1.69) |
| Plasma ALT, IU/L | 33 (25–45) | 28 (22–39) | 38 (30–50)*** | 30 (24–40) | 36 (27–48) |
| Plasma AST, IU/L | 30 (25–37) | 31 (25–38) | 30 (26–35) | 27 (24–32) | 33 (27–40)*** |
| AST/ALT ratio | 0.92 (0.76–1.10) | 1.03 (0.90–1.27) | 0.77 (0.70–0.98)*** | 0.90 (0.70–1.12) | 0.93 (0.77–1.10) |
| Plasma GGT, IU/L | 31 (22–46) | 28 (20–39) | 36 (25–52)* | 29 (21–44) | 32 (23–51) |
| Plasma ALP, IU/L | 65 ± 2 | 65 ± 3 | 66 ± 2 | 65 ± 2 | 66 ± 2 |
| Plasma albumin, g/L | 38 ± 1 | 38 ± 1 | 38 ± 1 | 38 ± 1 | 38 ± 1 |
| Platelet count, 10<sup>9</sup>/L | 242 (207–292) | 230 (207–285) | 258 (221–304) | 242 (204–299) | 244 (210–290) |
| Serum adiponectin, µg/mL | 7.8 (4.8–10.7) | 9.8 (6.2–12.3) | 6.0 (4.2–8.8)*** | 7.2 (4.8–9.9) | 8.4 (4.8–11.5) |
| IHTG content (histology), % | 10 (3–30) | 5 (0–25) | 20 (5–30)** | 5 (0–20) | 20 (5–30)** |
| Type 2 diabetes, *n* (%) | 50 (49) | 22 (43) | 28 (54) | 21 (42) | 29 (55) |
| Use of metformin, *n* (%) | 47 (46) | 21 (41) | 26 (50) | 18 (36) | 29 (55) |
| Use of statins, *n* (%) | 37 (36) | 18 (35) | 19 (37) | 18 (36) | 19 (36) |
| Cigarette smoking, *n* (%) | 23 (22) | 13 (26) | 10 (19) | 12 (24) | 11 (21) |
| Genetic risk score‡ | 0.27 ± 0.02 | 0.28 ± 0.03 | 0.25 ± 0.02 | 0.10 ± 0.01 | 0.43 ± 0.02*** |

Data are shown as *n* (%), mean ± SEM or median (25<sup>th</sup>–75<sup>th</sup> percentiles). Statistical tests used were the Student's *t*-test, Mann-Whitney *U* test, Pearson's $\chi^2$ test, or the Fisher's exact test, as appropriate. \**P* < 0.05; \*\**P* < 0.01; \*\*\**P* < 0.001 for 'high HOMA-IR' vs. 'low HOMA-IR' or 'high GRS' vs. 'low GRS'.
†Available for *n* = 91.
‡Calculated using the PNPLA3 rs738409, TM6SF2 rs58542926, MBOAT7 rs641738, and GCKR rs1260326 genotypes as described in Methods.

Abbreviations for all detected BAs are defined in **Table S1**. As expected, the liver contained a markedly higher proportion of primary BAs than serum (75.9 ± 1.24% *vs.* 41.0 ± 1.58%) (**Fig. 1B**). Almost the entire hepatic BA pool was conjugated (99.6 ± 0.02%), consisting of 76.3 ± 0.94% glycine conjugates and 23.7 ± 0.94% taurine conjugates (**Fig. 1C**). By contrast, only 45.9 ± 2.04% of serum BAs were conjugated, with glycine and taurine conjugates accounting for 88.1 ± 0.64% and 11.9 ± 0.64%, respectively (**Fig. 1C**). In serum, the most abundant individual BAs were the unconjugated and conjugated species of CDCA (a primary BA) and DCA (a secondary BA), which together comprised over 60% of the serum BA pool (**Fig. 1D**). On the other hand, the primary BAs CA and CDCA were the most abundant species in liver, comprising over 90% of the hepatic BA pool (**Fig. 1E**). Absolute and relative concentrations of all detected BAs are summarized in **Table S2**.

Most BAs in serum were significantly correlated with their levels in liver (**Fig. 1F**). Consistent with the established utility of circulating 7α-hydroxy-4-cholesten-3-one (C4) as a marker of hepatic BA biosynthesis,^36^ primary BAs in serum and particularly in liver showed strong correlations with serum C4 concentrations (**Fig. 1F**). Strikingly, most serum BAs had a highly positive association with plasma glucose, and both primary BAs as well as the secondary BAs tDCA and tUDCA correlated positively with serum insulin (**Fig. 1G**). These same BAs exhibited positive relationships with IHTG content, whereas only CDCA conjugates were significantly correlated with all histological features of MASLD, *i.e.*, steatosis, ballooning, inflammation, and fibrosis (**Fig. 1G**). In stark contrast, liver BAs had practically no associations with plasma glucose or serum insulin, liver histology, or circulating biomarkers of hepatic fibrogenesis (PRO-C3) or necroinflammation (CK-18 M65) (**Fig. 1H**). Hepatic primary BAs showed strong positive correlations with plasma triglycerides (**Fig. 1H**), a pattern which was mirrored by serum C4 concentrations (r = 0.53, *P* < 0.001; data not shown).

### A serum BA fingerprint identifies insulin-resistant individuals with high IHTG content

We first performed an unbiased clustering analysis of BA profiles to determine whether they delineate groups with distinct metabolic characteristics.

Unsupervised Bayesian model-based clustering based of serum BA concentrations resulted in three clusters of patients with unique BA signatures (C1, *n* = 37; C2, *n* = 32; C3, *n* = 34) (**Fig. 2A–B**). Low circulating BAs characterized C1, whereas C2 and C3 showed markedly greater total, primary, and secondary BAs (**Fig. 2C–E**). The BA increase in C2 was driven predominantly by unconjugated species (**Fig. 2F**), while in C3 the increase was mostly attributable to BA conjugates (**Fig. 2G**).

**Figure 2.**
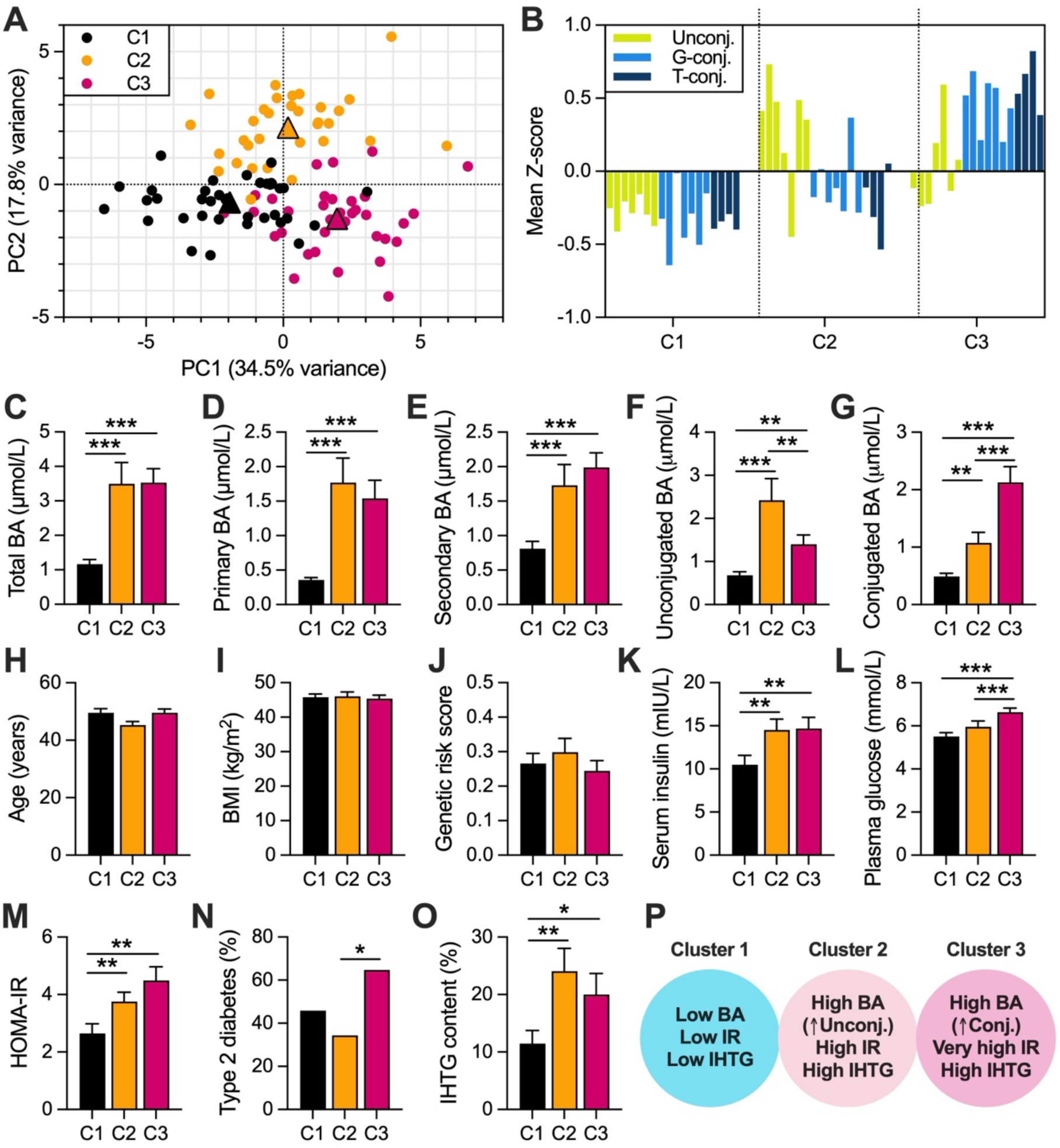
A serum BA fingerprint identifies individuals with insulin resistance and high IHTG content. (**A**) Principal component analysis (PCA) of serum BA concentrations. Individual subjects are shown as circles, color-coded based on model-based clustering (C1, black; C2, orange; C3, magenta). Triangles indicate the mean PC1/PC2 position of each cluster. (**B**) Mean Z-scores of standardized serum BA concentrations for the three clusters. Each bar represents a single BA, color-coded by conjugation state (unconjugated, green; glycine-conjugated, light blue; taurine-conjugated, dark blue). (**C**–**G**) Serum concentrations of (**C**) total BAs, (**D**) primary BAs, (**E**) secondary BAs, (**F**) unconjugated BAs, and (**G**) conjugated BAs in the clusters. (**H**–**O**) Clinical features of the clusters including (**H**) age, (**I**) BMI, (**J**) GRS, (**K**) serum insulin, (**L**) plasma glucose, (**M**) HOMA-IR, (**N**) prevalence of type 2 diabetes, and (**O**) IHTG content. (**P**) Summary of cluster characteristics. Data are shown as mean ± SEM. The Kruskal-Wallis test was used, with post hoc testing by the two-stage linear step-up procedure of Benjamini, Krieger, and Yekutieli. \**P* < 0.05; \*\**P* < 0.01; \*\*\**P* < 0.001. IR, insulin resistance.

Regarding clinical and metabolic characteristics, the clusters were similar with respect to sex distribution (63–71% females), age, BMI, and the GRS of IHTG content (**Fig. 2H–J**). However, compared with C1, serum insulin concentrations were significantly higher in C2 and C3 (**Fig. 2K**). Plasma glucose was also higher in C2 compared to C1 and in C3 compared to C2 (**Fig. 2L**). In accordance, HOMA-IR, a marker of hepatic insulin resistance, increased progressively from C1 to C3 (**Fig. 2M**). Consistent with marked insulin resistance in C3, this cluster had the highest prevalence of type 2 diabetes (**Fig. 2N**). Histological IHTG content was significantly higher in C2 and C3 compared to C1 (**Fig. 2O**). These findings indicate that elevated circulating BA concentrations, particularly BA conjugates, identify individuals with impaired insulin sensitivity and hepatic steatosis (**Fig. 2P**).

In contrast to serum BAs, clustering of the cohort based on liver BA concentrations did not yield biochemically or clinically meaningful groupings (data not shown).

### Circulating but not intrahepatic BAs increase in MASLD

We next divided the liver biopsy cohort into obese controls without steatosis (*n* = 26) and patients with MASLD (*n* = 77) (**Table S3**). MASLD was further categorized by the SAF score into mild (SAF < 3, *n* = 54) and moderate-to-severe (SAF ≥ 3, *n* = 23).

Hepatic BA concentrations did not differ between the groups, but MASLD induced major changes in circulating BAs (**Fig. 3A**) (**Table S4**). Total serum BAs increased from 1.6 ± 0.2 μmol/L in controls to 2.8 ± 0.3 μmol/L (+75%) in mild MASLD and 3.7 ± 0.8 μmol/L (+130%) in moderate-to-severe MASLD (*P*_trend_ < 0.001). This stepwise increase was clearly observed in primary, secondary, and conjugated BAs (**Fig. 3B–D**), whereas the increase in unconjugated BAs was borderline significant (**Fig. 3E**). Given that IHTG accumulation reflects both hepatic insulin resistance and genetic risk,^39^ we examined how these factors varied with MASLD severity. Mean HOMA-IR increased by 50% in mild MASLD and 110% in moderate-to-severe MASLD, while mean GRS increased by 40% and 90%, respectively (**Fig. 3F–G**).

**Figure 3.**
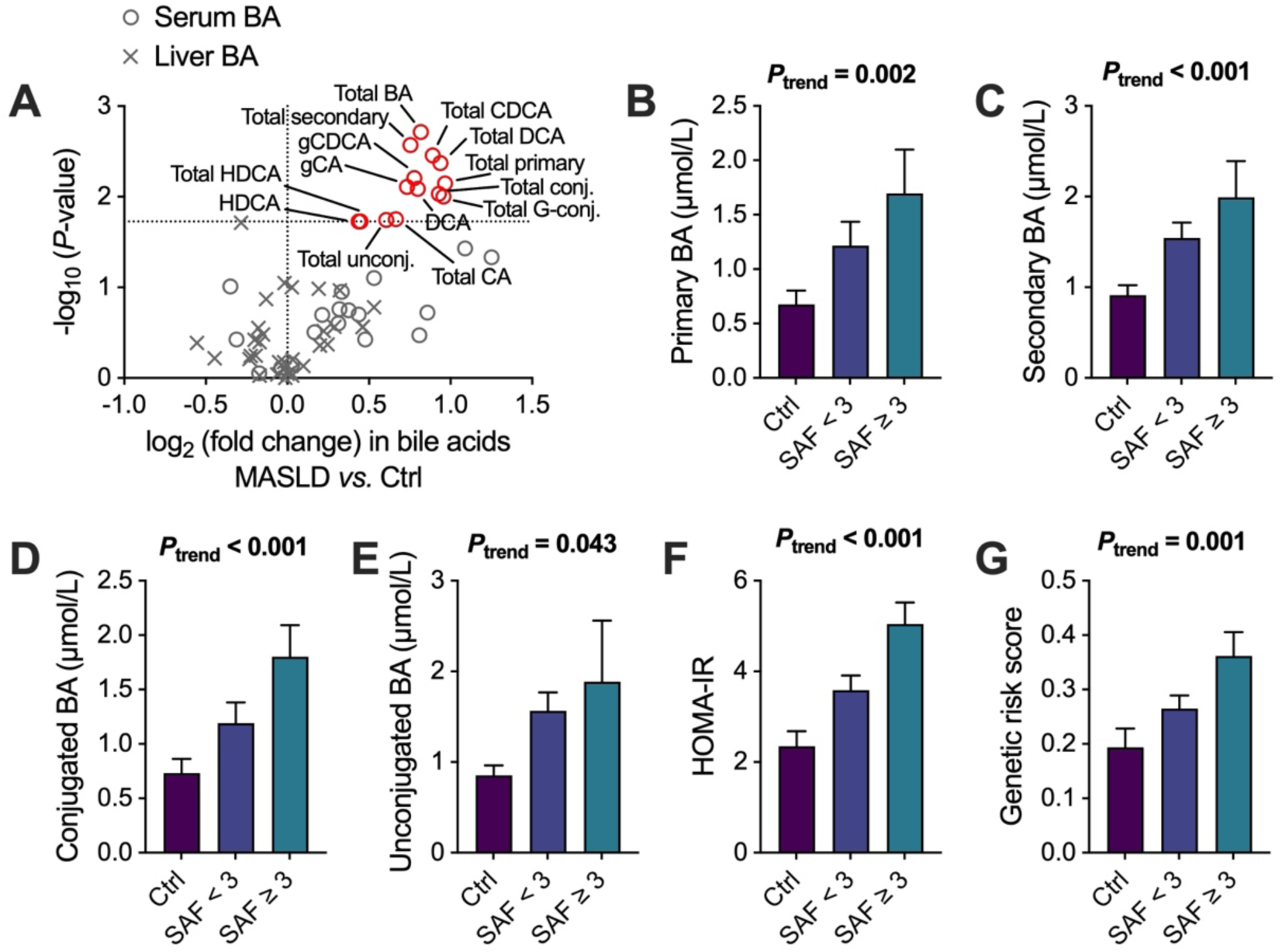
Circulating but not intrahepatic BAs increase in MASLD. (**A**) Volcano plot showing effects of MASLD on serum and liver BA concentrations. The x-axis denotes log_2_ fold change (MASLD *vs.* Ctrl), and the y-axis denotes −log_10_ *P*-value from the Mann-Whitney *U* test. Nominal *P*-values are shown, but the significance threshold (horizontal dotted line) was determined by the Benjamini-Hochberg method. (**B**–**G**) Comparisons of (**B**) primary BAs, (**C**) secondary BAs, (**D**) conjugated BAs, (**E**) unconjugated BAs, (**F**) HOMA-IR, and (**G**) GRS between controls and patients with mild (SAF < 3) or moderate-to-severe (SAF ≥ 3) MASLD. The Jonckheere-Terpstra test was used. Bar graph data are mean ± SEM.

Thus, the MASLD-related rise in circulating BAs occurred in parallel with increases in both insulin resistance and genetic susceptibility. This concordance makes it difficult to determine whether BA alterations reflect steatosis *per se* or the accompanying metabolic dysfunction.

### Insulin resistance, rather than IHTG, drives increases in circulating BAs

To disentangle the respective contributions of insulin resistance and IHTG to BA metabolism, we stratified the cohort using two orthogonal instruments: (1) HOMA-IR, an index of hepatic insulin resistance; and (2) a widely validated GRS for IHTG, which influences steatosis but not insulin sensitivity (**Fig. 4A; Table 1**).^4,5,7,39,40^ The GRS enables causal inference regarding the effect of steatosis on BAs, consistent with the principles of Mendelian randomization.^41^

**Figure 4.**
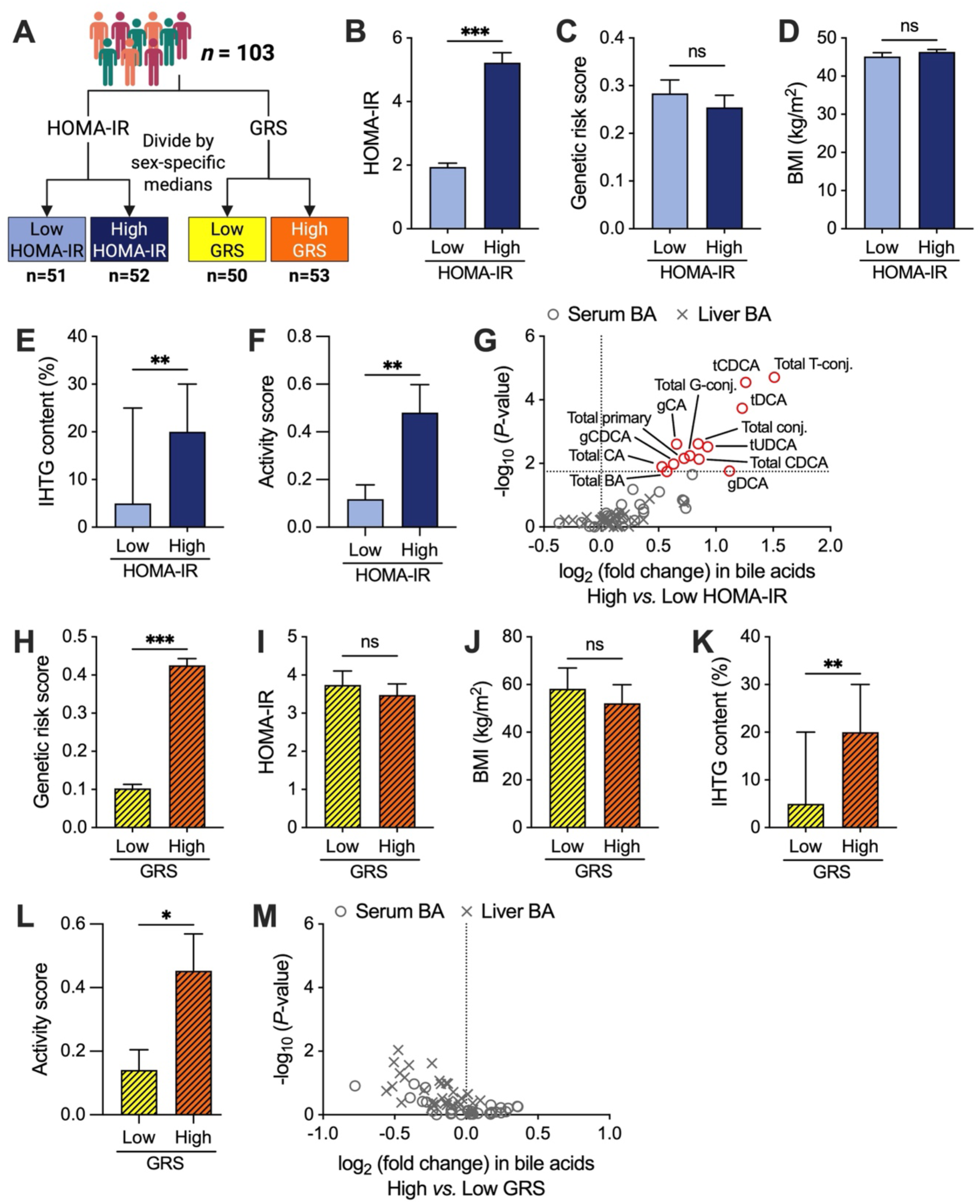
Insulin resistance and not IHTG drives MASLD-related increases in circulating BAs. (**A**) Study design. (**B**–**F**) Comparisons of (**B**) HOMA-IR, (**C**) GRS, (**D**) BMI, (**E**) IHTG content, and (**F**) SAF activity score between the low and high HOMA-IR groups. (**G**) Volcano plot showing effects of HOMA-IR on serum and liver BAs. The x-axis denotes log_2_ fold change (high *vs.* low HOMA-IR), and the y-axis denotes −log_10_ *P*-value. Nominal *P*-values are shown, but the significance threshold (horizontal dotted line) was determined by the Benjamini-Hochberg method. (**H**–**L**) Comparisons of (**H**) GRS, (**I**) HOMA-IR, (**J**) BMI, (**K**) IHTG content, and (**L**) SAF activity score between the low and high GRS groups. (**M**) Volcano plot showing effects of GRS on serum and liver BAs. The x-axis denotes log_2_ fold change (high *vs.* low GRS), and the y-axis denotes −log_10_ *P*-value. No Benjamini-Hochberg-adjusted *P*-values reached statistical significance. Bar graph data are mean ± SEM except for (**E**) and (**K**), which are median ± IQR. Mann-Whitney *U* test: \**P* < 0.05; \*\**P* < 0.01; \*\*\**P* < 0.001.

By definition, the high HOMA-IR group had a substantially greater mean HOMA-IR than the low HOMA-IR group (**Fig. 4B**), whereas GRS (**Fig. 4C**) and BMI (**Fig. 4D**) were similar. Histologically, the high HOMA-IR group exhibited a 4-fold greater median IHTG content and significantly higher activity scores (**Fig. 4E–F**). Multiple serum BA species were markedly elevated in the high versus low HOMA-IR group (**Fig. 4G; Table S5**), but hepatic BA concentrations remained unchanged (**Fig. 4G; Table S6**). Conversely, the high GRS group had a greater mean GRS than the low GRS group (**Fig. 4H**), while HOMA-IR (**Fig. 4I**) and BMI (**Fig. 4J**) were similar. Despite a comparable 4-fold increase in IHTG content and significantly higher activity scores (**Fig. 4K–L**), the high GRS group showed no differences in circulating or hepatic BA concentrations relative to the low GRS group (**Fig. 4M; Tables S5–S6**).

To address potential residual confounding, we performed in the full cohort multivariable linear regression relating BA concentrations to HOMA-IR and GRS, adjusting for age, sex, BMI, type 2 diabetes, smoking, and use of antidiabetic or lipid-lowering medications. HOMA-IR models were adjusted for the GRS and *vice versa*. The results were consistent with unadjusted group comparisons (**Tables S7–S8**). In sensitivity analyses, fibrosis stage and activity score did not independently predict BA concentrations after accounting for HOMA-IR (data not shown).

Finally, we confirmed in an independent liver biopsy cohort that genetically driven steatosis does not affect circulating BA levels. In his cohort, a positive association between HOMA-IR and plasma BAs has been shown previously.^35^ Participants were stratified by PNPLA3-I148M genotype into carriers (IM/MM, *n* = 20) and non-carriers (II, *n* = 36) (**Table S9**). The groups were similar in age, sex, HOMA-IR (**Fig. S1A**), and BMI (**Fig. S1B**). As expected, carriers displayed significantly higher histological steatosis scores (**Fig. S1C**) and more lobular inflammation (**Fig. S1D**) and ballooning (**Fig. S1E**). Despite this greater disease severity in variant carriers, circulating BAs did not differ between the groups (**Fig. S1F**; **Table S10**).

Taken together, our genetic approach demonstrates that MASLD-related increases in circulating BAs are driven by insulin resistance and cannot be attributed to IHTG accumulation.

### Hepatic insulin resistance increases circulating BAs without altering biosynthesis or transintestinal flux

We next characterized the pattern of BA elevation associated with insulin resistance within the main liver biopsy cohort. Individuals in the high HOMA-IR group had ∼60% greater total serum BA concentrations than those in the low HOMA-IR group (3.3 ± 0.5 *vs.* 2.0 ± 0.2 mmol/L; **Fig. 5A**). This was attributable to marked increases in both primary BAs (+106%) and secondary BAs (+35%) (**Fig. 5B–C**), specifically glycine- and taurine-conjugated species (**Fig. 5D–E**). Unconjugated BAs were not altered (*P* = 0.342; **Fig. 5F**). HOMA-IR was not associated with either the 12α-hydroxylated/non-12α-hydroxylated BA ratio (**Fig. 5G**) or the serum BA pool hydrophobicity index (**Fig. 5H**). However, a greater HOMA-IR was associated with reduced proportions of unconjugated and increased proportions of conjugated BAs (**Fig. 5I**). Partial least squares discriminant analysis (PLS-DA) identified the BAs most predictive of high HOMA-IR, in descending order: tCDCA, tDCA, gCA, tUDCA, gCDCA, and gDCA (**Fig. 5J**).

**Figure 5.**
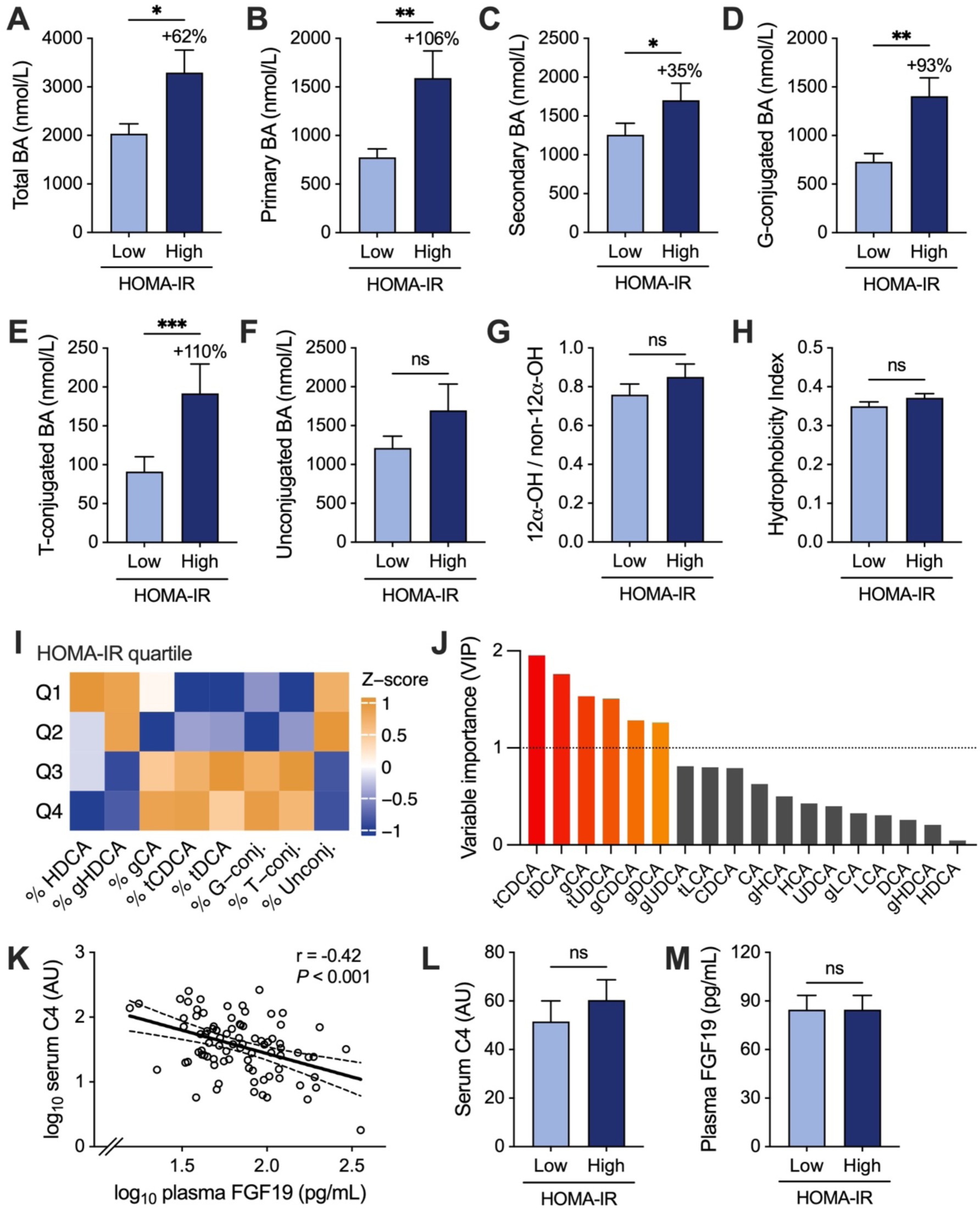
Hepatic insulin resistance increases circulating BA concentrations without alterations in BA biosynthesis or intestinal FGF19 signaling. (**A**–**H**) Comparisons of serum (**A**) total BAs, (**B**) primary BAs, (**C**) secondary BAs, (**D**) glycine-conjugated BAs, (**E**) taurine-conjugated BAs, (**F**) unconjugated BAs, (**G**) ratio of 12α-hydroxylated to non-12α-hydroxylated BAs, and (**H**) BA hydrophobicity index between the low and high HOMA-IR groups. (**I**) Heatmap illustrating standardized Z-scores of relative BA abundances across quartiles of HOMA-IR. Data are shown only for BAs with significantly (*P* < 0.05) altered relative abundances as a function of HOMA-IR. Blue color denotes lower and orange color higher Z-scores. (**J**) Variable importance in projection (VIP) scores for serum BAs derived from a multivariate PLS-DA model for the high *vs.* low HOMA-IR comparison. The horizontal dotted line marks the threshold (VIP > 1) above which BAs are considered to have a significant influence on HOMA-IR. (**K**) Relationship between log_10_-transformed concentrations of plasma FGF19 and serum C4. The line fit was obtained by linear regression. (**L**–**M**) Comparison of (**L**) serum C4 and (**M**) plasma FGF19 concentrations between the low and high HOMA-IR groups. Mann-Whitney *U* test: \**P* < 0.05; \*\**P* < 0.01; \*\*\**P* < 0.001.

In principle, elevated circulating BAs could reflect increased hepatic biosynthesis or enhanced intestinal absorption. Hepatic BA synthesis proceeds mainly through the classical pathway, for which C4 serves as a circulating marker (**Fig. 1A**).^36^ On the other hand, greater intestinal BA flux would be expected to raise FGF19 and suppress C4 via feedback inhibition of CYP7A1 (**Fig. 1A**).^37,38^ As anticipated, plasma FGF19 and serum C4 were inversely correlated (**Fig. 5K**). However, both C4 (**Fig. 5L**) and FGF19 (**Fig. 5M**) were similar between the high and low HOMA-IR groups. These data argue against altered hepatic BA synthesis or transintestinal flux as primary drivers of the elevated circulating BAs in insulin-resistant individuals.

### Fasting serum BAs are unaffected by overfeeding with fats or carbohydrates

We considered the possibility that habitual diet composition might underlie the elevation of fasting serum BAs in insulin-resistant individuals. To explore this, we conducted a post-hoc analysis of a previous overfeeding study in which 38 overweight participants were provided a surplus of 1000 kcal/day for three weeks, where the excess calories were derived from either saturated fat (SAT), unsaturated fat (UNSAT), or simple sugars (CARB).^42^ As previously described, all three regimens produced modest but statistically significant increases in IHTG content and a similar gain in body weight of 1.2 ± 0.2 kg (*P* < 0.001).^42^ However, fasting serum BA concentrations were unaffected by any of the dietary interventions (**Fig. S2**), indicating that even substantial differences in recent fat or sugar intake are unlikely to explain the association between HOMA-IR and serum BAs in the present study.

### The BA uptake transporter NTCP is downregulated in insulin-resistant livers

Next, we investigated whether hepatic insulin resistance influences the transcriptional regulation of genes involved in BA metabolism. To this end, the hepatic transcriptome was analyzed in two cohorts comprising patients undergoing a liver biopsy during metabolic surgery. Primary analysis was conducted in the discovery cohort (*n* = 127), followed by replication in the validation cohort (*n* = 86). **Table S11** shows characteristics of the cohorts.

Across the global transcriptome, 3,241 genes were differentially regulated as a function of IHTG content in the discovery cohort, with 1,865 transcripts downregulated and 1,376 upregulated (**Fig. 6A**). Focusing on manually curated gene sets related to several key aspects of BA metabolism (**Table S12**), we found a total of 30 differentially expressed genes involved in pathways of BA biosynthesis, conjugation and detoxification, signaling regulation, transport, and bile secretion (**Fig. 6B–F**). Of the 30 genes, 18 replicated in the validation cohort (**Fig. 6G**). Strikingly, clustering based on the expression profiles of these validated genes resulted in spontaneous segregation of the participants according to insulin sensitivity (**Fig. 6H**). Out of the 18 genes, 11 correlated significantly with HOMA-IR (**Fig. 6H**). Functional details for these genes are summarized in **Table 2**.

**Figure 6.**
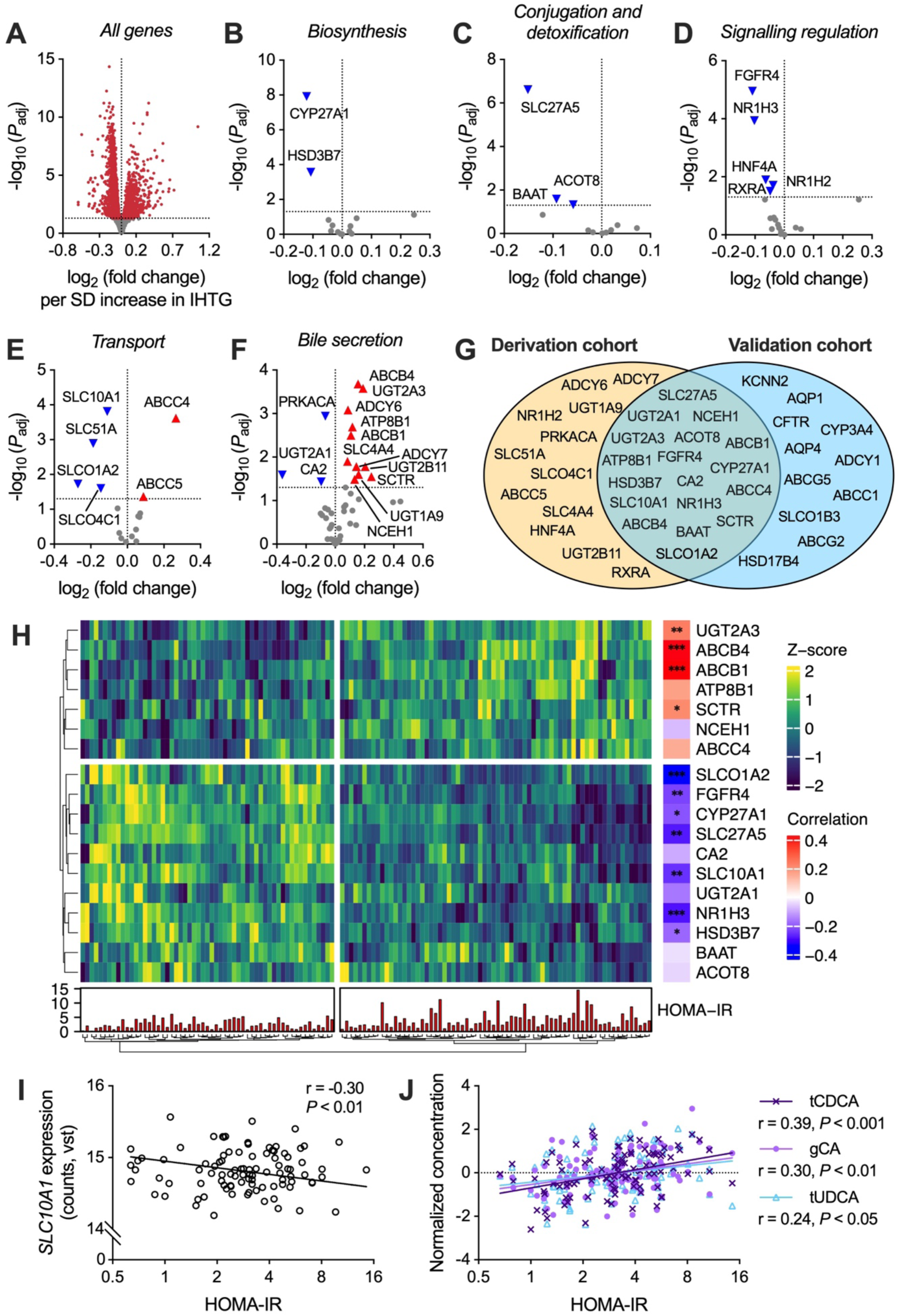
The BA uptake transporter NTCP is downregulated in insulin-resistant livers. (**A**–**F**) Differential expression analysis in the derivation cohort linking histological IHTG content to the hepatic transcriptome, adjusted for age and sex. Differentially expressed genes are shown for (**A**) all genes (*n* = 16,449) and for pathways involved in (**B**) BA biosynthesis, (**C**) BA conjugation and detoxification, (**D**) signaling regulation of BA metabolism, (**E**) BA transport, and (**F**) bile secretion. The x-axis denotes log_2_ fold change in gene expression per 1 SD increase in IHTG, and the y-axis denotes −log_10_ adjusted *P*-value. Significantly up- or downregulated genes are indicated by labeled triangles. (**G**) Venn diagram of differentially regulated BA metabolism-related genes in the derivation and validation cohorts. (**H**) Heatmap showing standardized (Z-score) expression of differentially regulated genes in the derivation and validation cohorts as a function of IHTG content. Rows represent genes, and columns represent individual subjects. A secondary heatmap on the right shows Pearson correlation coefficients between gene expression and HOMA-IR, and the lower bar plot shows HOMA-IR values for each subject. \**P* < 0.05; \*\**P* < 0.01; \*\*\**P* < 0.001. (**I**) Relationship between HOMA-IR and normalized expression of *SLC10A1*/NTCP. (**J**) Relationships between HOMA-IR and normalized serum concentrations of tCDCA (dark purple crosses), gCA (light purple circles), and tUDCA (light blue triangles).

**Table 2.** Validated BA metabolism-related which were differentially regulated by IHTG and correlated with HOMA-IR.

| Gene symbol | Direction | Protein product | Function | Refs. |
| --- | --- | --- | --- | --- |
| *Biosynthesis* |  |  |  |  |
| CYP27A1 | Down | Sterol 27-hydroxylase | Initiating enzyme of the alternative/acidic pathway of BA synthesis. | 44,45 |
| HSD3B7 | Down | 3β-hydroxysteroid dehydrogenase type 7 | Converts 7α-hydroxycholesterol to C4 as the second step of the classical BA biosynthetic pathway. | 45 |
| *Conjugation and detoxification* |  |  |  |  |
| SLC27A5 | Down | Long-chain fatty acid transport protein 5 (FATP5; formerly bile acyl-CoA synthetase [BACS]) | Catalyzes the formation of BA-CoA thioesters for subsequent conjugation with glycine or taurine; transports long-chain fatty acids into hepatocytes. | 46 |
| *Signaling regulation* |  |  |  |  |
| FGFR4 | Down | Fibroblast growth factor receptor 4 | FGF19 receptor; activation represses CYP7A1 and inhibits BA synthesis (part of the FXR-FGF19 gut-liver axis). | 47 |
| NR1H3 | Down | Oxysterols receptor LXR-α (also liver X receptor alpha [LXR-α]) | Oxysterol-activated nuclear receptor; promotes cholesterol efflux and lipogenesis; induces CYP7A1 in mice but not in humans. | 48 |
| *Transport* |  |  |  |  |
| SLC10A1 | Down | Hepatic sodium/bile acid cotransporter (also sodium/taurocholate co-transporting polypeptide [NTCP]) | Basolateral Na<sup>+</sup>-dependent uptake of conjugated BAs. Principal hepatocellular BA uptake transporter; receptor for hepatitis B/D viruses. | 49 |
| SLCO1A2 | Down | Solute carrier organic anion transporter family member 1A2 (OATP1A2) | Uptake transporter for amphipathic organic anions and bile salts; in liver, apical on cholangiocytes. | 50 |
| *Bile secretion* |  |  |  |  |
| UGT2A3 | Up | UDP-glucuronosyltransferase 2A3 | BA glucuronidation in human liver, at least DCA, CDCA, UDCA, and HDCA. | 51 |
| SCTR | Up | Secretin receptor | Increases bicarbonate secretion in bile ducts. | 52 |
| ABCB4 | Up | PC translocator ABCB4 (also multidrug resistance protein 3 [MDR3]) | Floppase actively translocating PC from the inner to the outer leaflet of the hepatocellular canalicular membrane, mediating biliary phospholipid secretion. | 53 |
| ABCB1 | Up | ATP-dependent translocase ABCB1 (also multidrug resistance protein 1 [MDR1] or P-glycoprotein 1 [P-gp]) | Polyspecific efflux pump on the canalicular membrane; mediates biliary export of many hydrophobic/amphipathic xenobiotics and drugs. | 54 |

We found that hepatic SLC10A1 was markedly downregulated as a function of IHTG content (**Fig. 6E** and **6H**), and its expression was inversely correlated with HOMA-IR (**Fig. 6I**). This gene encodes NTCP, which is the principal BA uptake transporter on the hepatocyte sinusoidal membrane.^43^ NTCP has a high affinity for conjugates of both dihydroxylated (CDCA, DCA, UDCA) and trihydroxylated (CA) BAs,^43^ which emerged among the strongest predictors of hepatic insulin resistance in the present study (**Fig. 5J**). Accordingly, serum levels of these BAs showed associations with HOMA-IR in the opposite direction to hepatic SLC10A1/NTCP expression (**Fig. 6J**). These findings implicate reduced NTCP-mediated hepatic uptake as a potential driver of elevated circulating BAs in insulin resistance.

### Insulin acutely suppresses the splanchnic spillover of BA conjugates

We next investigated whether insulin acutely modulates systemic BA spillover from the splanchnic circulation. To this end, we measured BAs in paired arterial and hepatic venous blood samples in five male subjects with biopsy-confirmed MASLD who underwent hepatic venous catheterization (mean age 51 ± 5 years; mean BMI 28.1 ± 4.3 kg/m^2^).^55^ Samples were obtained basally and during euglycemic hyperinsulinemia.

Under basal conditions, concentrations of individual BA species did not differ between arterial and hepatic venous blood (data not shown). During hyperinsulinemia, conjugated BAs decreased markedly, with similar 30–40% reductions in arterial and hepatic venous blood (**Fig. 7A**), whereas unconjugated BAs were unaffected (**Fig. 7B**). The insulin-mediated suppression involved both primary and secondary BA conjugates, which decreased to comparable extents (**Fig. 7C**). All individual conjugated BA species showed a tendency to decrease, with significant reductions in tDCA, gCDCA, tCDCA, and gCA (**Fig. 7D**). Although not statistically significant, the degree of suppression was inversely related to IHTG content, with attenuated responses in individuals with more severe steatosis (**Fig. 7E**). Collectively, these findings support the hypothesis that insulin acutely increases splanchnic clearance of BA conjugates, thereby limiting their spillover into the systemic circulation.

**Figure 7.**
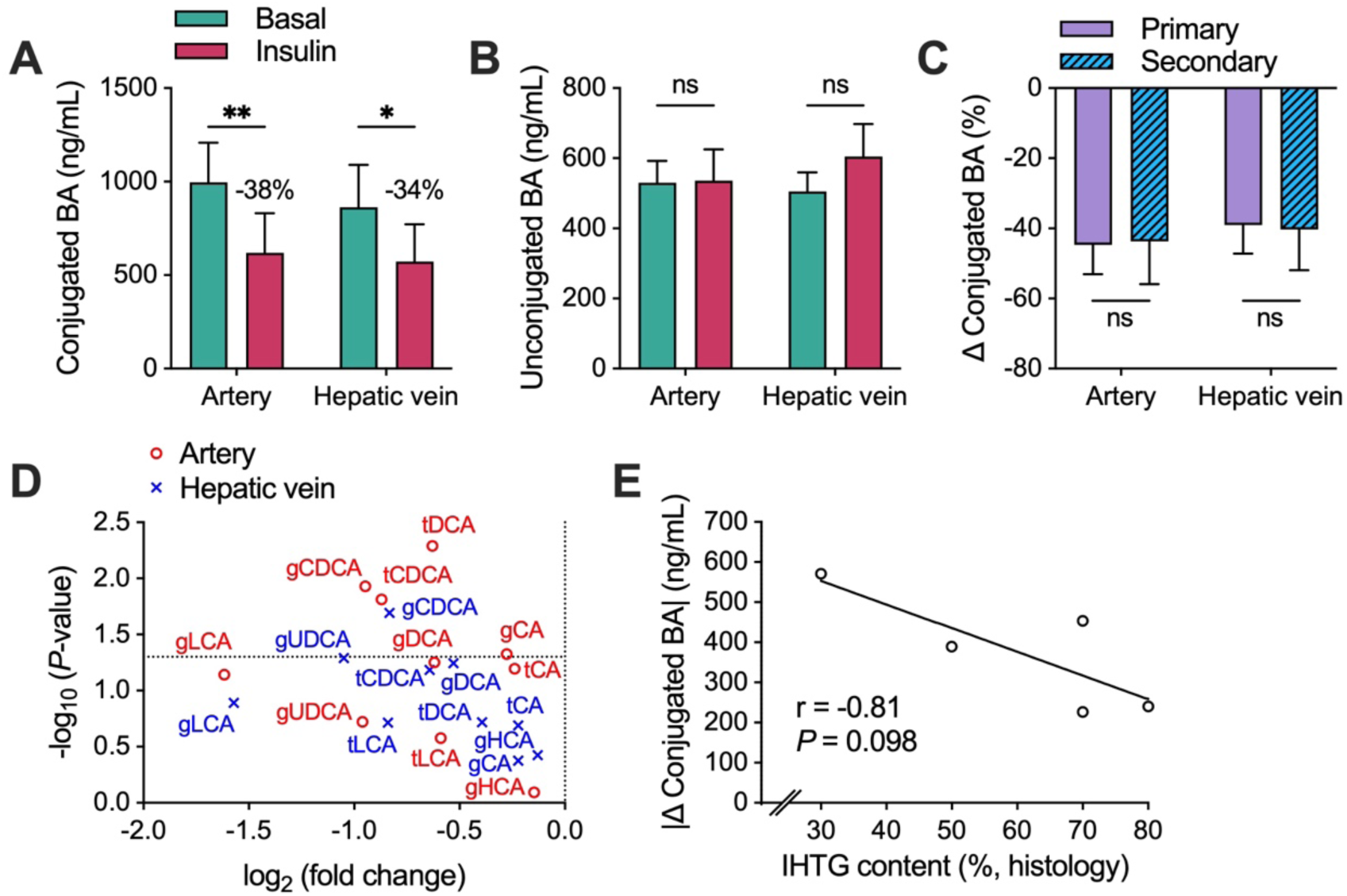
Insulin acutely suppresses splanchnic spillover of BA conjugates. (**A**–**B**) Concentrations of (**A**) conjugated and (**B**) unconjugated BAs in arterial and hepatic venous blood under basal conditions and during euglycemic hyperinsulinemia. (**C**) Percent change in primary and secondary conjugated BAs during insulin infusion. Paired *t*-test: \**P* < 0.05; \*\**P* < 0.01. (**D**) Volcano plot showing effects of hyperinsulinemia on individual conjugated BA concentrations. The x-axis denotes log_2_ fold change (hyperinsulinemia *vs.* basal), and the y-axis denotes −log_10_ *P*-value from the paired *t*-test. (**E**) Relationship between IHTG content and absolute change in plasma conjugated BA concentration during insulin infusion. The line fit was obtained by linear regression.

### Impaired hepatic clearance explains elevated circulating BAs in a physiology-based simulation of human liver metabolism

Finally, to evaluate whether impaired hepatic BA clearance could account for the elevated circulating BAs observed in insulin-resistant individuals, we used a recently developed physiologically based pharmacokinetic (PBPK) model of human BA metabolism.^56^ Stepwise reductions in hepatic uptake of conjugated BAs produced proportional increases in 24-hour plasma BA concentrations, while intrahepatic BA levels remained unchanged (**Fig. S3A**). In the fasted state (08:00 AM), decreasing hepatic clearance to 75% and 50% of normal raised total plasma BAs by ∼20% and ∼60%, respectively (**Fig. S3B**), driven by ∼30% and ∼100% increases in conjugated BAs (**Fig. S3C**). Unconjugated BAs were unaffected (**Fig. S3D**). Fasting plasma FGF19, C4, and the hepatic BA synthesis rate were likewise unaltered (**Fig. S3E–G**). These simulations mirror our findings *in vivo* and support impaired hepatic BA clearance as a plausible mechanism for the observed increases in circulating BAs.

## Discussion

Bile acid dysmetabolism is a recognized feature of MASLD, yet its mechanistic basis has remained unclear.^18^ By leveraging validated genetic instruments of IHTG content to enable causal inference, we found that BA abnormalities in MASLD arise from insulin resistance rather than hepatic steatosis. Circulating – but not intrahepatic – BAs increased in parallel with impaired glycemia and hepatic insulin sensitivity, accompanied by reduced expression of the hepatic BA uptake transporter NTCP. We also provide evidence that insulin may directly modulate splanchnic BA spillover into the systemic circulation.

Hepatic and circulating BAs were highly interrelated (**Fig. 1**), which is remarkable given the functional and anatomical disconnect between these two pools: only a minor fraction of hepatocellular BAs undergo sinusoidal efflux, while biliary BAs must first traverse the intestine before potentially entering the systemic circulation.^10^ This is likely explainable by efficiency of the enterohepatic circulation, which renders each BA pool interdependent by nature. Serum C4 correlated highly with primary BAs, not only in liver but also in serum, indicating that alterations in BA biosynthesis are directly reflected in circulating BAs. Hepatic BAs have rarely been quantified in human studies, but where comparative data are available, our measurements align with previously published serum and liver BA pool compositions.^19,20,57–63^

Circulating BAs provided a metabolomic signature that robustly identified patients with severe insulin resistance and MASLD, independent of age, BMI, or genetic predisposition (**Fig. 2**). Accordingly, serum BAs correlated strongly with glucose, insulin, HbA1c, and both circulating and histological markers of liver injury (**Fig. 1**), reinforcing data from previous reports.^18,34^ MASLD itself associated with a doubling of serum BAs, with stepwise increases as a function of disease severity (**Fig. 3**). However, these BA elevations closely paralleled increases in both HOMA-IR and GRS, reflecting the metabolic and genetic drivers of IHTG accumulation. This demonstrates that insulin resistance inherently confounds the association between BAs and MASLD when dividing the cohort based on histological diagnosis.

Genetic instruments have been essential in delineating causal relationships between MASLD and its metabolic correlates.^64^ Using HOMA-IR and a validated GRS for IHTG, we separated the effects of insulin resistance from those of steatosis (**Fig. 4**). Individuals with high *vs.* low HOMA-IR showed marked increases in both serum BAs and hepatic insulin resistance, while having a similar GRS. In contrast, the high *vs.* low GRS groups showed no differences in insulin sensitivity or BA concentrations. Because IHTGs increased 4-fold in both comparisons, these data point to insulin resistance, rather than steatosis itself, as the driver of BA elevations. This interpretation was reinforced in an external cohort using the PNPLA3-I148M variant (**Fig. S1**). In that cohort, Legry *et al.* demonstrated that circulating BAs do not differ between MASH patients and BMI- and HOMA-IR-matched controls, but increase only in those with severe insulin resistance.^35^ Subsequently, Grzych *et al.* found that plasma BAs are elevated specifically in insulin-resistant patients with MASH.^34^ The pattern of BA changes observed in high *vs.* low HOMA-IR – greater increases in conjugated than unconjugated BAs and in primary than secondary BAs (**Fig. 5**) – mirrors previous studies relating BAs with insulin resistance, type 2 diabetes, and MASLD.^18^ Although direct comparisons are limited by differences in study design, our finding of a ∼60% increase in total serum BAs in insulin-resistant subjects aligns with previous estimates ranging from 34% to 75% in analogous comparisons.^23,26,35,65^ Contrary to a previous report, we saw no changes in the 12α-hydroxylated/non-12α-hydroxylated BA ratio or in the BA hydrophobicity index.^23^

Intrahepatic BAs did not correlate with metabolic or histological features of MASLD (**Fig. 1; Fig. 3**) and were unchanged as a function of insulin resistance or genetic risk (**Fig. 4**). This was unexpected given the markedly increased serum BAs in insulin resistance, as well as the prevailing view that cytotoxic BA species play a role in MASH pathogenesis.^14–17^ We are aware of two prior studies which have quantified hepatic BAs in human MASLD. Aranha *et al.* reported increased DCA but no other differences in 15 MASH patients *vs.* 8 healthy liver donors.^19^ Lake *et al.*, analyzing post-mortem or other biopsied liver samples without detailed clinical characterization, described heterogeneous BA changes in MASH (*n* = 37) *vs.* healthy (*n* = 17) or steatotic (*n* = 7) livers, including decreased CA and gDCA and increased tCA, tDCA, and gCDCA.^20^ Both studies were limited by small sample size, variable tissue sources, insufficient phenotyping, and uncertain pre-analytical conditions, complicating interpretation. In contrast, our data provide no evidence that intrahepatic BA levels are significantly altered in insulin resistance or MASLD without advanced fibrosis.

How does insulin resistance lead to higher circulating BAs? Portal venous blood delivers gut-absorbed BAs to the liver, where 50–90% are extracted depending on species.^10^ The fraction that escapes hepatic clearance spills over into the systemic circulation. Thus, serum BA levels reflect net balance between intestinal absorption and hepatic clearance.^56,66^ In MASLD, elevated serum BAs have been ascribed to enhanced biosynthesis, inferred from increased hepatic expression of the rate-limiting CYP7A1 enzyme.^30,67^ We found no evidence supporting this, as CYP7A1 expression was unchanged in two cohorts, with no alterations in intrahepatic BAs or serum C4 (**Fig. 5–6**). Intestinal BA flux also appeared unaffected, as FGF19 levels were unchanged in subjects with high HOMA-IR (**Fig. 5**). Furthermore, dietary influences were unlikely to explain these findings, as three weeks of fat overfeeding failed to alter fasting serum BA concentrations (**Fig. S2**). Taken together, these data suggest that the BA dysmetabolism observed in insulin resistance is not driven by increased biosynthesis, altered intestinal transport, or dietary factors, but rather by another as-yet unidentified mechanism.

A more plausible explanation is impaired hepatic clearance of portal BAs, which is primarily mediated by the basolateral transporter NTCP.^43^ In two RNA-seq cohorts, NTCP expression was inversely associated with both IHTG and HOMA-IR (**Fig. 6**), consistent with prior reports of reduced NTCP mRNA^15,68,69^ and protein levels^70–72^ in human MASLD. NTCP preferentially transports conjugates of CDCA, DCA, CA, and UDCA,^43^ which were the species most strongly associated with HOMA-IR in our study (**Fig. 5**). Circulating concentrations of these same BA conjugates increase markedly following pharmacological NTCP inhibition with bulevirtide in humans.^73^ Notably, the positive associations between these conjugated BAs and HOMA-IR mirrored the inverse relationship between HOMA-IR and NTCP expression (**Fig. 6**). Together, these findings implicate reduced hepatic BA uptake as the mechanistic basis for elevated circulating BAs in insulin-resistant states.

No studies have demonstrated a direct role for insulin in regulating hepatic BA clearance. We explored this by analyzing BAs in samples from a previous study that combined hepatic vein catheterization with a hyperinsulinemic-euglycemic clamp.^55^ Surprisingly, a 120-minute insulin infusion led to a marked suppression of conjugated BAs in both arterial and hepatic venous blood, whereas unconjugated BAs were unaltered (**Fig. 7**). Unconjugated BAs are poor NTCP substrates, and their hepatic uptake involves alternative mechanisms.^10,43^ Although the small sample size limited statistical power, the suppression of conjugated BAs tended to be weaker in individuals with higher IHTG content. Consistent with these observations, Haeusler *et al.* reported a 44% insulin-mediated decrease in serum BAs in lean individuals, with attenuation of this effect in obesity.^22^ These data support the hypothesis that insulin may modulate hepatic BA clearance, and that this putative action exhibits features of insulin resistance. They also align with predictions from a human-based PBPK model showing that reduced hepatic BA clearance is sufficient to reproduce the phenotype seen in insulin resistance *in vivo* (**Fig. S3**).

A substantial body of evidence supports BAs as important regulators of systemic energy homeostasis.^8^ Nevertheless, it remains uncertain whether elevations in circulating BAs could exert physiologically meaningful effects. In mice, NTCP inhibition with bulevirtide increases BA concentrations and improves metabolic parameters, including reductions in body weight and IHTG content and enhanced glucagon-like peptide-1 secretion.^74^ Such effects are not readily apparent in humans,^75^ although most studies of bulevirtide have focused on treatment of viral hepatitis. Clinical trials of FXR agonists and FGF19 mimetics consistently reduce IHTG and improve MASH,^76^ but these effects are thought to arise from direct hepatic signaling and are therefore not easily reconcilable with findings from the present study. It remains to be confirmed whether serum BAs could serve as actionable biomarkers of metabolic dysfunction or predictors of therapeutic response to BA-modulating interventions.

To the best of our knowledge, this is the first study to quantify both circulating and hepatic BAs in a large cohort spanning the full clinical spectrum of IHTG content. A major strength was the use of the GRS to separate the effects of insulin resistance from those of IHTG. Human-relevant mechanistic insight was obtained via post-hoc analyses of prior interventional studies. No participants had advanced fibrosis, which would be a potential confounder due to accompanying cholestasis.^31,77^ This also means that our data do not exclude a role for BAs in liver injury at later disease stages. A limitation is that all measurements were obtained in the fasted state, preventing assessment of postprandial and diurnal BA dynamics.^78^ Further, BAs in liver biopsy homogenates reflect a mixture of intrahepatocellular and canalicular pools, whose relative contributions cannot be distinguished. Finally, the hepatic vein catheterization study, while informative, was limited by small sample size and the absence of a saline control.

In conclusion, our findings argue against a major pathogenic role for BAs in MASLD but uncover a previously unrecognized link between circulating BA dynamics and insulin action. Advances in analytical chemistry have markedly expanded the BA landscape, revealing hundreds of new molecular variants with unknown physiological functions.^79^ Future work should confirm the role of insulin signaling in hepatic BA clearance and define the clinical significance of disturbances within this increasingly complex metabolic network.

## Data availability

The datasets generated and/or analyzed during the present study are not publicly available but are available from the corresponding author upon reasonable request.

## Author contributions

Conceptualization, S.F.Q.; investigation, S.F.Q., S.J., K.P., A.J., A.K.P., J.A., E.V., M.O., T.T., T.H.; formal analysis, S.F.Q.; validation, J.T.H., E.D., L.V., S.F., B.S.; writing—original draft, S.F.Q.; writing—review & editing, S.F.Q., J.T.H., B.S., H.Y.-J.; visualization, S.F.Q.; project administration, S.F.Q.; resources, H.Y-J.; supervision, S.Q., H.Y-J.

## Acknowledgements

The authors gratefully acknowledge the volunteers for their help. We thank Päivi Ihamuotila and Aila Karioja-Kallio for excellent technical assistance; Bo Angelin, Ingela Arvidsson, and Jennifer Härdfeldt for valuable discussions on data interpretation; and Veronika Voronova for advice on the PBPK model. This study was supported by the Orion Research Foundation (S.F.Q.); the Yrjö Jahnsson Foundation (20207313 to S.F.Q.); the Maud Kuistila Memorial Foundation (2021-0301B to S.F.Q.); the Emil Aaltonen Foundation (210182 to S.F.Q.); the Finnish Medical Foundation (5843 to S.F.Q.); the Biomedicum Helsinki Foundation (20230241 to S.F.Q.); the Academy of Finland (309263 to H.Y.-J.; 315589 and 320129 to T.T.); the Finnish Government special state subsidy (EVO) granted to HUS Helsinki University Hospital (H.Y-J.); the Sigrid Jusélius Foundation (H.Y.-J.; T.T.); the Novo Nordisk Foundation (NNF19OC0057503 to H.Y.-J.); the French ANR: LabEx EGID (10-LABX-0046), RHU PreciNASH (16-RHUS-0006), and DeCodeNASH (ANR-20-CE14-0034); an ERC Advanced Grant (694717 to B.S.); an ERC Starting Grant (101042759 to J.T.H.); HEPADIP (Hepatic and adipose tissue and functions in the metabolic syndrome, EU 6th Framework Program, LSHM-CT-2005-018734); and RESOLVE (A systems biology approach to RESOLVE the molecular pathology of two hallmarks of patients with metabolic syndrome and its co-morbidities, hypertriglyceridemia and low HDL-cholesterol, EU 7th Framework Program [Grant agreement no. 305707]).

## Materials and Methods

### Subjects and study design

These studies complied with the Declaration of Helsinki. All participants provided a written informed consent after being explained the nature and potential risks of the procedures.

### Main liver biopsy cohort and liver RNA-seq cohorts

For analysis of serum and liver BAs, we consecutively recruited 103 subjects undergoing a liver biopsy during metabolic surgery at HUS Helsinki University Hospital (Helsinki, Finland). This cohort is denoted the main liver biopsy cohort. Liver transcriptomics were further analyzed in two separate cohorts of 127 and 86 subjects, each studied using similar procedures.

All subjects fulfilled the following inclusion criteria: (*i*) age between 18 and 75 years; (*ii*) no known acute or chronic illnesses other than obesity, type 2 diabetes, or hypertension, based on medical history, physical examination, electrocardiogram, and standard laboratory tests (complete blood count and serum creatinine, electrolyte, and thyrotropin concentrations); (*iii*) no clinical or biochemical signs of liver disease other than MASLD, such as chronic hepatitis B or C, hemochromatosis, Wilson’s disease, alpha-1 antitrypsin deficiency, autoimmune hepatitis, or cholestatic liver disease; (*iv*) no signs of inborn errors of metabolism; (*v*) alcohol consumption <20 g/day for women and <30 g/day for men; (*vi*) no history of exposure to medications or toxins that influence hepatic steatosis; and (*vii*) not pregnant or lactating.

The subjects underwent an intraoperative liver biopsy as described below, and part of the biopsy specimen was used for either BA profiling or RNA sequencing. A week prior to the date of surgery, the subjects attended a metabolic study visit following an overnight fast. A detailed medical history was obtained and a physical examination was performed. Body weight was measured to the nearest 0.1 kg using a calibrated digital scale (Soehnle, Backnang, Germany), with subjects wearing light indoor clothing and no shoes. Height was measured to the nearest 0.5 cm using a non-stretchable tape measure. Waist circumference was measured at the midpoint between the lower rib margin and the superior iliac spine, and hip circumference was measured at the level of the greater trochanter. Blood pressure was measured in the sitting position after a minimum of 15 min of acclimatization and prior to blood sampling using an automatic oscillometric sphygmomanometer (OMRON M7; Omron Healthcare, Kyoto, Japan). Blood samples were drawn from an antecubital vein for measurement of blood counts and concentrations of glucose, HbA1c, total cholesterol, HDL cholesterol, LDL cholesterol, triglycerides, ALT, AST, ALP, GGT, albumin, insulin, C-peptide, adiponectin, C4, FGF19 and BAs. An aliquot of whole blood was obtained for extraction of genomic DNA and genotyping of IHTG-associated genetic variants. After basal blood sampling, a 2-hour oral glucose tolerance test (OGTT) using 75 g of glucose was performed (*n* = 91 in the main liver biopsy cohort).

The study protocol was approved by the Ethical Review Committee of the Hospital District of Helsinki and Uusimaa.

### Antwerp validation cohort

The validation cohort included 56 subjects receiving care at the obesity clinic of the Antwerp University Hospital (Edegem, Belgium) in whom fasting plasma concentrations of BAs were measured. All subjects had overweight or obesity and underwent a liver biopsy for evaluation of MASLD. The study design has been described previously.^35^ Inclusion criteria were: (*i*) suspected MASLD based on elevated liver enzyme activities (ALT, AST, or GGT) or a finding of steatosis on ultrasound; (*ii*) absence of significant alcohol consumption (>20 g/day); (*iii*) no history of bariatric surgery; (*iv*) no clinical or biochemical evidence of chronic liver disease other than MASLD; (*v*) no prior diagnosis of type 2 diabetes; and (*vi*) no current use of anti-diabetic or lipid-lowering medications, or antibiotics.

Fasting blood samples were obtained for the quantification of plasma BA concentrations using high-performance liquid chromatography-tandem mass spectrometry (HPLC-MS/MS), and for measurements of glucose, HbA1c, insulin, lipids, and liver enzymes.^80,81^ Genotyping of PNPLA3-I148M was performed using TaqMan assays (Applied Biosystems Inc., Foster City, CA) on a LightCycler 480 real-time polymerase chain reaction (PCR) system (Roche, Penzberg, Germany). Liver histology was evaluated by two independent, experienced pathologists blinded to the subjects’ clinical data, according to the MASH Clinical Research Network framework.^82^ A diagnosis of MASH was made when steatosis, lobular inflammation, and hepatocellular ballooning were simultaneously present.^83^

The study protocol was approved by the Ethical Review Committee of the Antwerp University Hospital.

### Overfeeding cohort

The cohort comprised 38 overweight individuals who were recruited by newspaper advertisements or by contacting those who had previously participated in metabolic studies. All subjects met the general inclusion criteria described above, and additional study-specific exclusion criteria were: (*i*) diabetes type 1 or 2; (*ii*) BMI ≥ 40 kg/m^2^; and (*iii*) use of anti-diabetic or lipid-lowering medications. The study has been previously described in detail (NCT02133144; ClinicalTrials.gov).^42^ In brief, the subjects were randomly assigned to one of three dietary intervention groups, each providing a surplus of 1000 kcal/day for 3 weeks. The excess energy was provided either as saturated fat (SAT; *n* = 14), unsaturated fat (UNSAT; *n* = 12), or simple sugars (CARB; *n* = 12). Serum BA concentrations were measured at baseline and following the intervention period.

The study protocol was approved by the Ethical Review Committee of the Hospital District of Helsinki and Uusimaa.

### Hepatic vein catheterization cohort

We measured fasting BA concentrations in blood samples collected from five subjects who underwent a liver biopsy and hepatic venous catheterization at Karolinska Hospital (Stockholm, Sweden). The study originally comprised nine subjects who were referred to the gastroenterologist for evaluation of abnormal liver function tests. For this post-hoc analysis, we excluded four subjects: one due to insufficient sample volume, and three due to advanced liver fibrosis which can influence BA metabolism.^31^ The study has been previously described in detail.^55^ Briefly, a catheter was inserted into a right hepatic vein under fluoroscopic guidance. Following a 90 min basal infusion of normal saline, a primed-continuous insulin infusion (0.5 mIU/kg/min) was initiated for 120 min to induce euglycemic hyperinsulinemia. Plasma glucose was maintained at 5 mmol/L using a variable-rate 20% glucose infusion. Splanchnic blood flow was quantified using indocyanine green, infused at 0.04 mg/kg/min from 30 to 210 min. BA concentrations were analyzed in paired arterialized and hepatic venous plasma samples collected at 90 min (saline infusion) and at 210 min (hyperinsulinemia).

The study protocol was approved by the Ethical Review Committee of the Karolinska Hospital.

### Liver biopsies and histological assessment

A liver biopsy was performed at the beginning of the laparoscopic surgery procedure. Part of the biopsy was used for histological analysis, and the rest was immediately snap-frozen in liquid nitrogen for subsequent measurement of BA concentrations and analysis of the hepatic transcriptome. An expert liver pathologist performed central reading of the liver biopsies, blinded to the subjects’ identities and histories. Histopathological features of MASLD (steatosis, lobular inflammation, ballooning, fibrosis) were assessed using the Steatosis, Activity, Fibrosis (SAF) grading and definitions.^84^ The activity score (A0–A4) was calculated as the sum of scores of lobular inflammation (I0–I2) and ballooning (B0–B2). The total SAF score (ranging from 0 to 11) was calculated as the sum of scores of steatosis (S0–S3), activity (A0–A4), and fibrosis (F0–F4). We diagnosed NASH when steatosis, lobular inflammation, and ballooning were concomitantly present.^83^

### Measures of insulin sensitivity

As the primary measure of hepatic insulin resistance, we used fasting serum insulin and plasma glucose concentrations to calculate HOMA-IR using the formula: HOMA-IR = serum insulin (mIU/L) × plasma glucose (mmol/L) ÷ 22.5.^85^ The Matsuda insulin sensitivity index (ISI) was used as an additional measure of insulin resistance in the subjects who underwent OGTT, calculated based on plasma glucose and serum insulin concentrations measured at 0, 30, and 120 min after ingesting the glucose solution.^86^

### Genotyping and calculation of the GRS for IHTG content

All subjects were genotyped for IHTG-associated risk variants in PNPLA3 rs738409, TM6SF2 rs58542926, MBOAT7 rs641738, and GCKR rs1260326. Genomic DNA was extracted from whole blood, and approximately 10 ng of DNA was used for genotyping via the TaqMan PCR method following the manufacturer’s protocol (Applied Biosystems, Foster City, CA). Post-PCR allelic discrimination was performed by detecting allele-specific fluorescence using the ABI Prism Sequence Detection System ABI 7900HT (Applied Biosystems). Genotyping success rate was >95%. All genotype frequencies were consistent with Hardy-Weinberg equilibrium (*P* > 0.05 by Haldane exact test). These genotypes were used to calculate a previously validated GRS of IHTG content using the following formula: GRS = 0.266 × PNPLA3 rs738409:G (0/1/2) + 0.274 × TM6SF2 rs58542926:T (0/1/2) + 0.063 × MBOAT7 rs641738:T (0/1/2) + 0.065 × GCKR rs1260326:T (0/1/2).^4,5,7^

### Serum and liver BA profiling

Serum BA concentrations were quantified using ultra-high performance liquid chromatography coupled with quadrupole time-of-flight mass spectrometry (UHPLC-QTOF-MS), following a previously established sample preparation protocol.^87^ A 25 mg Ostro Protein Precipitation and Phospholipid Removal 96-well plate was used (Waters Corporation, Milford, MA). Each well received 100 mL of serum and 10 mL of an internal BA standard solution (1000 ng/mL), followed by 450 mL of acetonitrile containing 1% formic acid. Eluates were collected, dried under nitrogen, and reconstituted in 30 mL methanol, 10 mL BA recovery standard (1000 ng/mL), and 50 mL of 2 mM ammonium acetate in water. Chromatographic separation was performed on an ACQUITY UPLC® BEH C18 column (2.1 × 100 mm, 1.7 mm particle size; Waters) using an Agilent UHPLC-Q-TOF-MS system (Agilent Technologies, Santa Clara, CA). The mobile phases consisted of H_2_O:MeOH (70:30, v/v) for phase A and pure MeOH for phase B, both containing 2 mM ammonium acetate. The elution gradient was as follows: 5–30% B (0–1.5 min), 30–70% B (1.5–4.5 min), 70–100% B (4.5–7.5 min), holding at 100% B for 5.5 min, followed by a 5 min re-equilibration. Flow rate was 0.4 mL/min, with a total runtime of 18 min per sample. Ionization was achieved using a dual ESI source with the following parameters: capillary voltage 4.5 kV, nozzle voltage 1500 V, nebulizer N_2_ pressure at 21 psi, N_2_ sheath gas flow rate 11 L/min and temperature 379 °C. Data acquisition was performed in negative ion mode over m/z 100–1700. MassHunter B.06.01 (Agilent Technologies) was used for acquisition, and data were processed with MZmine 2.52.^88^ Quality control included blanks, pure standards, and plasma control samples. Identification of unknown compounds relied on exact mass and MS/MS fragmentation data.

Hepatic BA concentrations were determined from frozen liver samples following homogenization with a CryoPrep CP02. (Covaris, Woburn, MA) Approximately 10 mg of tissue was weighed, mixed with 12.5 mL of an internal standard solution and 0.5 mL of extraction solvent (5% ammonium hydroxide in acetonitrile), and homogenized for 2 min at 20 Hz using a mixer mill (MM301; Retsch GmbH, Haan, Germany). The supernatant was collected, and the pellet was subjected to a second extraction. Combined supernatants were dried under nitrogen, and the residue was reconstituted in 30 mL of 40% methanol. Subsequent steps followed previously described protocols.^89^ Analyses were carried out using a Waters Xevo TQ-S mass spectrometer coupled to an Acquity UPLC system (Waters, Milford, MA) equipped with an HSS T3 column (100 mm × 2.1 mm i.d., 1.7 mm particle size) and gradient elution. The assay targeted 33 known BAs and monitored 28 additional unidentified species (see ^89^). Quantification of targeted BAs was performed using the internal standard method, whereas semi-quantitative estimates of unidentified BAs were derived from calibration against the corresponding targeted BA and internal standard.

The relative hydrophilic-hydrophobic profile of BAs was assessed by calculating the bile salt monomeric hydrophobicity index according to the method of Heuman.^90^For each subject, the index was determined using the equation: HI = ∑*^n^* HI_x_F_x_, where HI_x_ represents the hydrophobicity index of the individual bile salt x; F_x_ is its mole fraction within the mixture; and *n* denotes the total number of distinct bile salts present.

### Liver transcriptomics

RNA was isolated from liver biopsy specimens using the AllPrep DNA/RNA/miRNA Universal Kit (QIAGEN, Hilden, Germany). Illumina Stranded mRNA Prep was used for construction of libraries (Illumina, San Diego, CA), and bulk RNA sequencing was performed on an Illumina platform, generating paired-end reads of 150 bp with an average depth of approximately 100 million reads per sample before filtering. Data processing was largely based on the GTEx V8 RNA-seq analysis workflow,^91^ with minor adaptations. Following quality control and trimming of adapter sequences, read alignment to the GRCh38/hg38 human reference genome was performed using STAR version 2.6.0a,^92^ with gene annotation according to GENCODE 26. Gene-level expression was calculated based on a collapsed gene model, where all isoforms were represented by a single transcript per gene, and read counts were generated via RNA-SeQC version 2.0.3.^93^

### Physiology-based PBPK model of human BA metabolism

Simulations were conducted to examine how reduced hepatic BA clearance influences plasma and liver BA concentrations, BA biosynthesis, and signaling. For this purpose, we applied a recently published PBPK model of human BA metabolism.^56^ This multi-compartment model employs a system of ordinary differential equations to describe production, transport, biotransformation, and elimination of CA, CDCA, and DCA in conjugated and unconjugated forms throughout the enterohepatic circulation, incorporating the FXR-FGF19 signaling axis. The model was constructed and parameterized using human physiological and experimental data and subsequently validated against published findings from diverse perturbations and pathologies.^56^

The PBPK model was originally implemented in Simulx (MonolixSuite; Simulations Plus, Research Triangle, NC) and subsequently adapted for the R package RxODE. The base simulation was run using default parameters.^56^ To simulate impaired hepatic clearance of BA conjugates, additional models were generated in which hepatic uptake rate constants were reduced to 75% and 50% of normal. These adjustments were intended to approximate diminished hepatic NTCP function, which was not explicitly represented in the model but is the principal transporter mediating the uptake of BA conjugates.^12,13^ All models were run to steady state, after which the final 24 hours of data were extracted. To enable comparison with clinical studies, analyses were focused on the 08:00 AM fasting time point. Extracted outputs included plasma and liver BA concentrations, plasma FGF19 and C4 levels, and rates of *de novo* BA synthesis.

### Other analytical procedures

Plasma glucose was measured using the hexokinase method on an autoanalyzer (Roche Diagnostics Hitachi 917; Hitachi Ltd., Tokyo, Japan). Serum insulin and C-peptide concentrations were determined by time-resolved fluoroimmunoassay using their respective kits (AutoDELFIA; Wallac, Turku, Finland). HbA1c was measured by HPLC using the fully automated Glycosylated Hemoglobin Analyzer System (Bio-Rad, Richmond, CA). Plasma triglycerides, total cholesterol, HDL cholesterol, and LDL cholesterol concentrations were measured using enzymatic kits; plasma ALT, AST, GGT, and ALP activities were measured using photometric International Federation of Clinical Chemistry methods; and plasma albumin was measured using a photometric method, on an autoanalyzer (Roche Diagnostics Hitachi 917). Complete blood counts were obtained using impedance, flow cytometry, and photometric detection (XN10; Sysmex, Kobe, Japan). Technicians blinded to clinical data used ELISAs to measure concentrations of serum adiponectin (Human Adiponectin ELISA; B-Bridge International Inc., San Jose, CA), serum PRO-C3 (Nordic Bioscience A/S, Herlev, Denmark), serum CK-18 fragments using the M65 antibody (M65 ELISA; VLVbio AB, Nacka, Sweden), and plasma FGF19 (Human FGF-19 Quantikine ELISA, DF1900; R&D Systems, Minneapolis, MN), with all analyses performed in duplicate.

### Statistical analyses

Analyses were performed in R version 4.2.1 (R Foundation for Statistical Computing, Vienna, Austria) and GraphPad Prism version 9.4.1 (GraphPad Software, La Jolla, CA) for macOS. Continuous variables were inspected for normality using the Shapiro-Wilk and/or Kolmogorov-Smirnov tests and Q-Q plots. Categorical variables are summarized as *n* (%) and continuous variables as mean ± SEM or median (25^th^–75^th^ percentiles), as appropriate.

For two independent groups, continuous variables were compared using the unpaired Student’s *t*-test (normally distributed data) or the Mann-Whitney *U* test (non-normally distributed data). Comparisons among three groups employed the Kruskal-Wallis test, with post-hoc testing via the two-stage linear step-up procedure of Benjamini, Krieger, and Yekutieli.^94^ Alternatively, ordered group differences were assessed with the exact Jonckheere-Terpstra trend test (R package clinfun), using 200,000 permutations. Categorical variables were compared using the Pearson’s ×^2^ test or Fisher’s exact test. For within-subject comparisons of continuous variables, a paired *t*-test was used. The nominal level of significance was α = 0.05. For analyses including a high number of multiple comparisons, false discovery rate was controlled using the Benjamini-Hochberg procedure. Unless otherwise indicated, unadjusted *P*-values are shown for results meeting discovery criteria.

We performed unbiased model-based clustering with finite Gaussian mixture models to identify groups of subjects with distinct BA profiles, as implemented in the R package mclust. The BA data were log-transformed and standardized to Z-scores. Clustering was run separately for serum and liver BAs using the Mclust function, with model selection based on the Bayesian Information Criterion. PCA was used for cluster visualization, and mean Z-scores of individual BA species were plotted to depict inter-cluster differences.

To investigate in the main liver biopsy cohort whether hepatic steatosis is causal in BA dysmetabolism, we leveraged a GRS of IHTG content. Because genetic variants are randomly allocated at conception, their effects are not subject to confounding, enabling causal inference between IHTG and BA metabolism through the principles of Mendelian randomization.^41^ To assess the impact of insulin resistance on BAs, we used HOMA-IR. For statistical analysis, the cohort was divided in two ways based on sex-specific median values of the GRS (males 0.298; females 0.266) and HOMA-IR (males 4.12; females 3.05). Subjects with GRS/HOMA-IR greater than or equal to the median were categorized as having high GRS/HOMA-IR, while those with GRS/HOMA-IR below the median were classified as having low GRS/HOMA-IR.

Multiple linear regression was used to analyze associations between HOMA-IR, GRS, and BA concentrations, with models adjusted for age, sex, BMI, type 2 diabetes, smoking status, and use of anti-diabetic or lipid-lowering medications, which were considered potential confounders of BA metabolism. Model diagnostics (residual, Q-Q, and scale-location plots) were used to verify linearity, residual normality, and homoscedasticity; no influential outliers were detected. To identify serum BAs best discriminating the high *vs.* low HOMA-IR groups, PLS-DA was conducted using the R package mixOmics. VIP scores >1 were considered as strong predictors.^95^

Differential expression analysis of the liver RNA-seq data was performed with the R package DESeq2 by using negative binomial generalized linear models.^96^ Histological IHTG content was the primary variable of interest, with age and sex as covariates, and continuous covariates were scaled to zero mean and unit variance. Pseudogenes were excluded and transcripts with <10 total reads were filtered out. Log_2_ fold changes were shrunk using the apeglm method.^97^ Significance was defined as Benjamini-Hochberg-adjusted *P*_adj_ < 0.05 after independent filtering on the mean of normalized counts. Quality control via principal component analysis, count heatmaps, and hierarchical clustering with clinical metadata showed no evidence of batch effects or outliers. Transcript-level downstream analyses used counts scaled by DESeq2-estimated size factors after variance stabilizing transformation.

## Supplementary Figures

**Figure S1.**
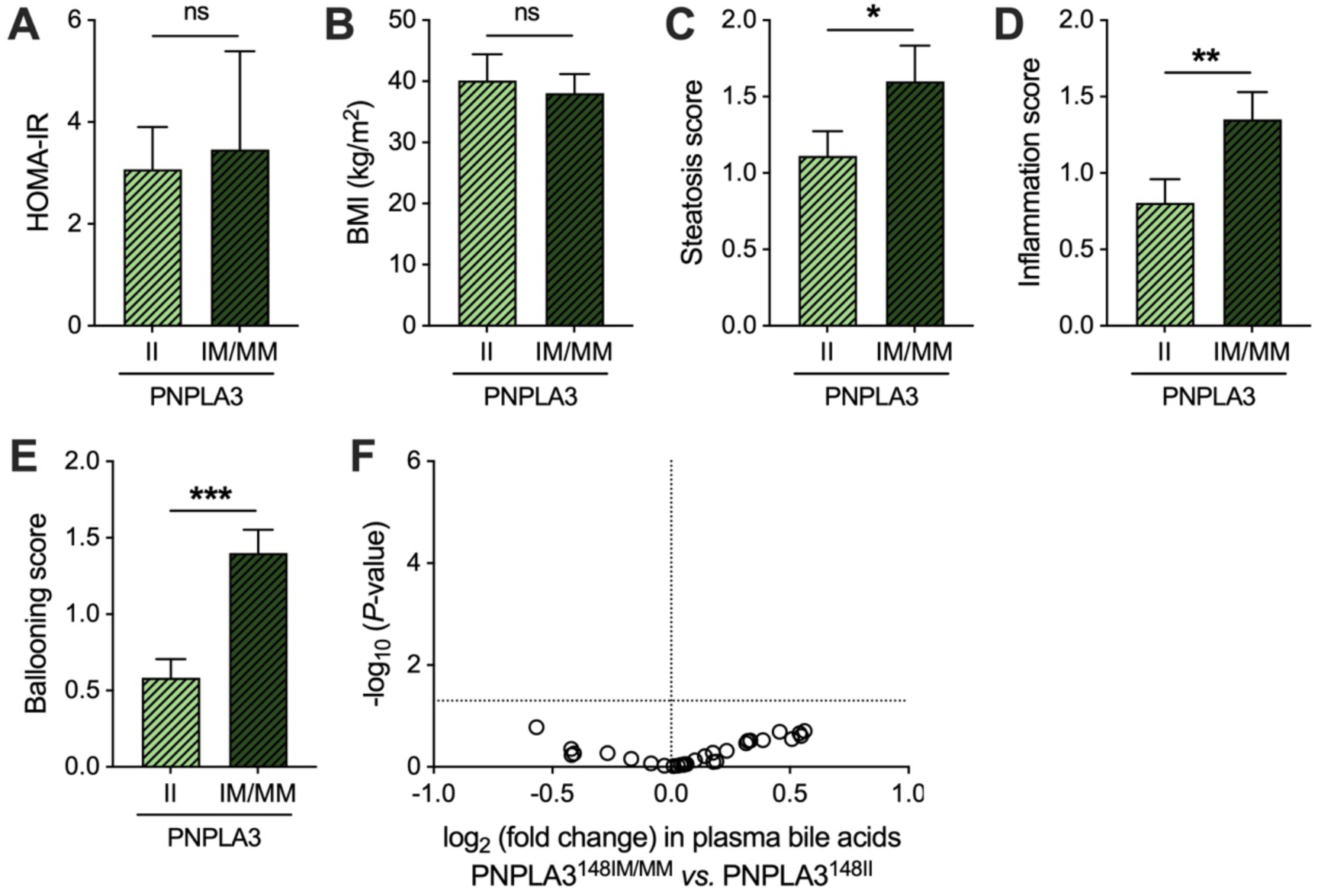
Genetic increase in IHTG content does not affect circulating BAs in an external validation cohort. (**A**–**E**) Comparisons of (**A**) HOMA-IR, (**B**) BMI, (**C**) histological steatosis score, (**D**) histological inflammation score, and (**E**) histological ballooning score between PNPLA3-I148M variant carriers and non-carriers. (**F**) Volcano plot showing effects of PNPLA3-I148M on plasma BA concentrations. The x-axis denotes log_2_ fold change (PNPLA3^148IM/MM^ *vs.* PNPLA3^148II^), and the y-axis denotes −log_10_ *P*-value. Bar graph data are mean ± SEM. The Mann-Whitney *U* test was used for all comparisons. \**P* < 0.05; \*\**P* < 0.01; \*\*\**P* < 0.001.

**Figure S2.**
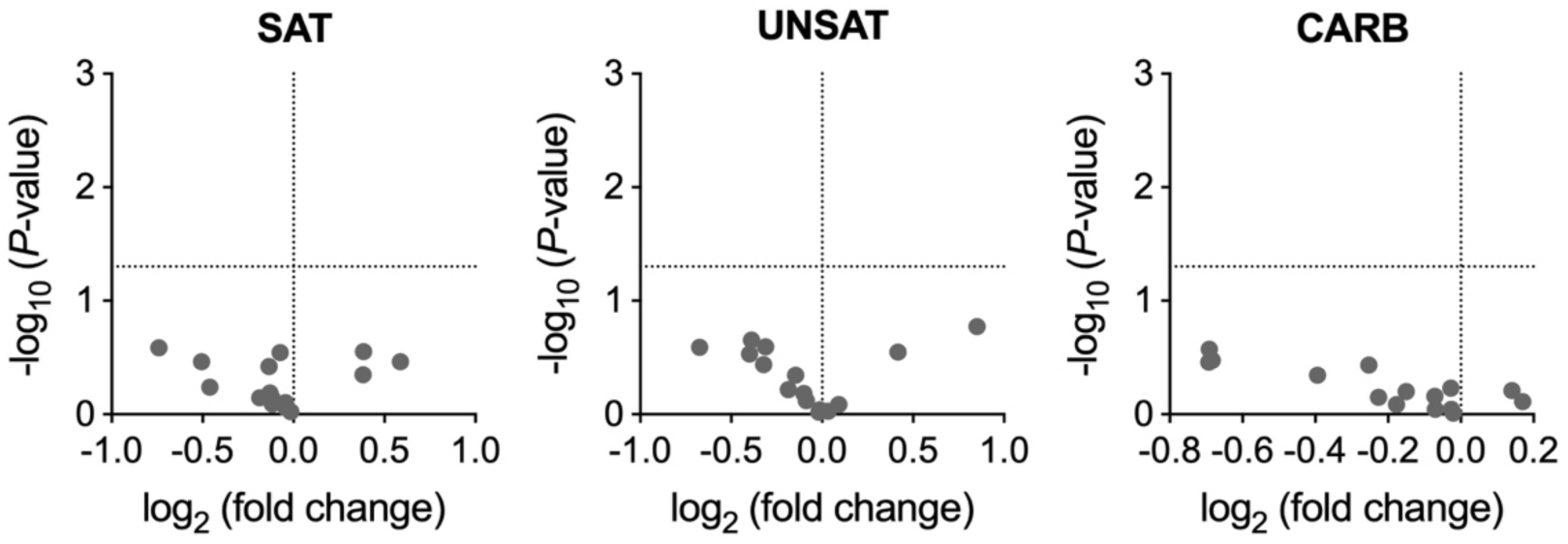
Fasting serum BAs are unaffected by overfeeding with fats or carbohydrates. Volcano plots showing effects of overfeeding on serum BA concentrations in the SAT (saturated fat), UNSAT (unsaturated fat), and CARB (simple sugars) groups. The x-axis denotes log_2_ fold change in BAs after the intervention compared to baseline, while the y-axis denotes −log_10_ *P*-value from the paired *t*-test.

**Figure S3.**
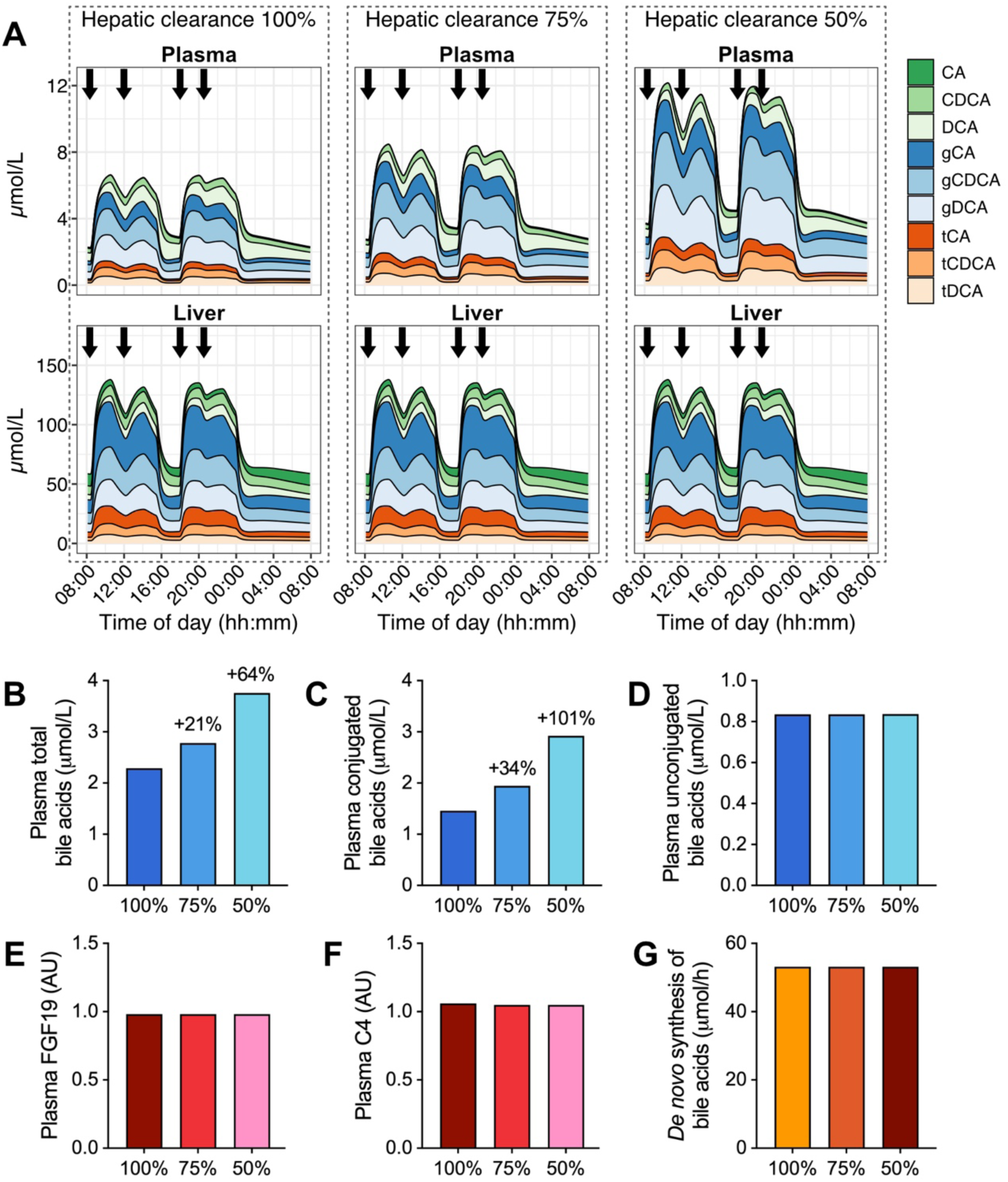
Impaired hepatic clearance explains elevated circulating BAs in a physiology-based model of human liver metabolism. **(A)** Simulated 24-hour BA concentrations within plasma and liver compartments, showing the effects of stepwise reduced hepatic clearance of conjugated BAs from the sinusoidal space: 100% (left), 75% (middle), and 50% (right). Meals were administered at 08:30, 12:00, 18:00, and 20:30 in the simulations (arrows). **(B–G)** Model outputs for plasma concentrations of **(B)** total BAs, **(C)** conjugated BAs, **(D)** unconjugated BAs, **(E)** FGF19, and **(F)** C4, as well as **(G)** hepatic *de novo* BA synthesis rate, simulated at 100%, 75%, and 50% clearance. All parameters were evaluated at *t* = 08:00, corresponding to the overnight-fasted state.

## Supplementary Tables

**Table S1.**
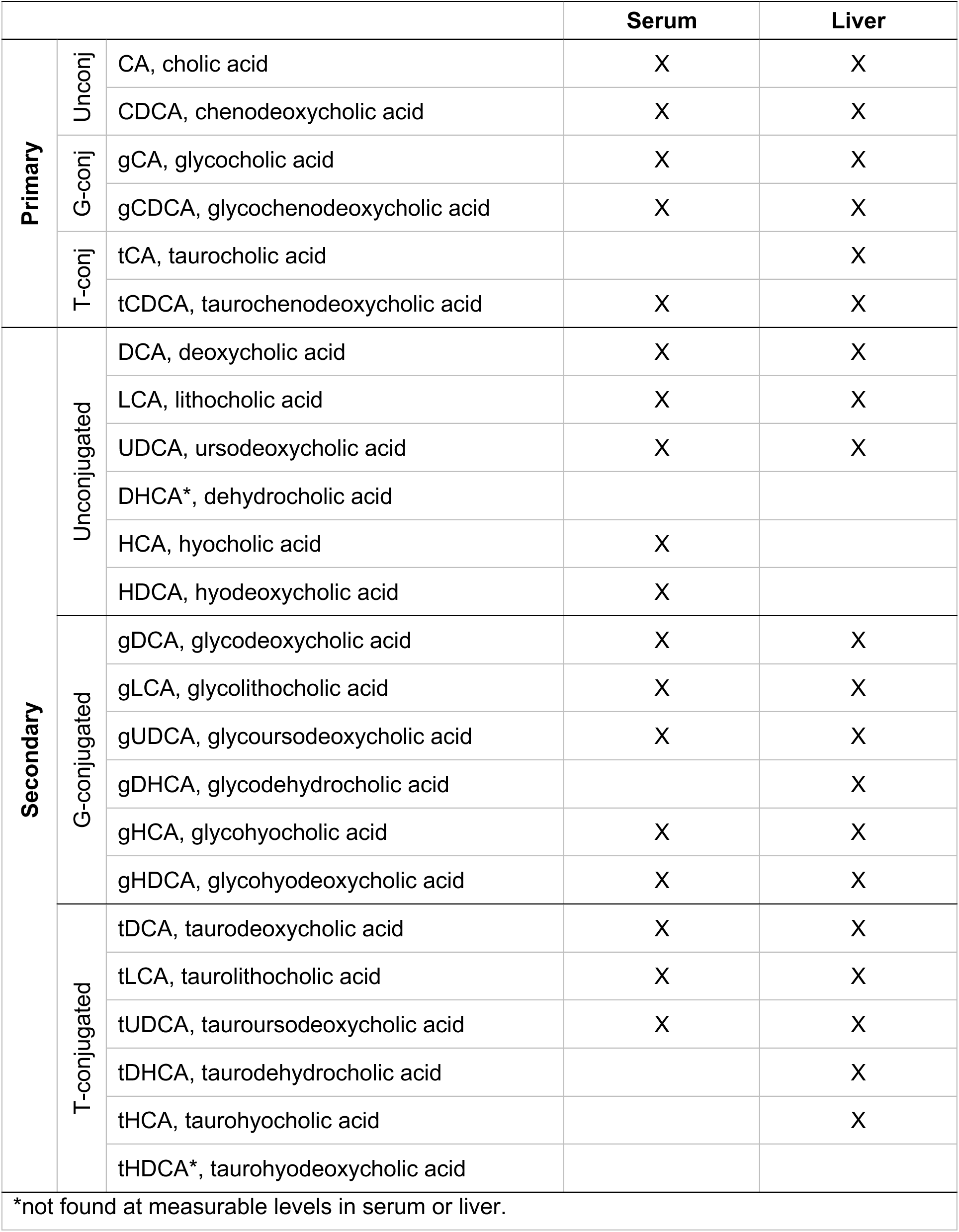
Detected bile acid species in serum and liver.

|  |  |  | Serum | Liver |
| --- | --- | --- | --- | --- |
| Primary | Unconj | CA, cholic acid | X | X |
|  |  | CDCA, chenodeoxycholic acid | X | X |
|  | G-conj | gCA, glycocholic acid | X | X |
|  |  | gCDCA, glycochenodeoxycholic acid | X | X |
|  | T-conj | tCA, taurocholic acid |  | X |
|  |  | tCDCA, taurochenodeoxycholic acid | X | X |
| Secondary | Unconjugated | DCA, deoxycholic acid | X | X |
|  |  | LCA, lithocholic acid | X | X |
|  |  | UDCA, ursodeoxycholic acid | X | X |
|  |  | DHCA*, dehydrocholic acid |  |  |
|  |  | HCA, hyocholic acid | X |  |
|  |  | HDCA, hyodeoxycholic acid | X |  |
|  | G-conjugated | gDCA, glycodeoxycholic acid | X | X |
|  |  | gLCA, glycolithocholic acid | X | X |
|  |  | gUDCA, glyoursodeoxycholic acid | X | X |
|  |  | gDHCA, glycodehydrocholic acid |  | X |
|  |  | gHCA, glycohyocholic acid | X | X |
|  |  | gHDCA, glycohyodeoxycholic acid | X | X |
|  | T-conjugated | tDCA, taurodeoxycholic acid | X | X |
|  |  | tLCA, taurolithocholic acid | X | X |
|  |  | tUDCA, tauroursodeoxycholic acid | X | X |
|  |  | tDHCA, taurodehydrocholic acid |  | X |
|  |  | tHCA, taurohyocholic acid |  | X |
|  |  | tHDCA*, taurohyodeoxycholic acid |  |  |
| *not found at measurable levels in serum or liver. |  |  |  |  |

**Table S2.** Absolute and relative concentrations of BAs in the main liver biopsy cohort.

|  | Serum, nmol/L |  | Liver, nmol/g × 10 |  |
| --- | --- | --- | --- | --- |
|  | Median [IQR] | % (Mean ± SD) | Median [IQR] | % (Mean ± SD) |
| Total CA | 110.37 [218.84] | 9.51 ± 7.55 | 211.43 [190.59] | 38.72 ± 10.26 |
| CA | 24.34 [39.59] | 3.66 ± 6.03 | 0.12 [0.09] | 0.02 ± 0.01 |
| gCA | 71.44 [116.69] | 5.85 ± 5.11 | 165.42 [143.85] | 29.24 ± 8.79 |
| tCA | ND | ND | 47.98 [38.81] | 9.46 ± 4.85 |
| Total CDCA | 568.53 [817.41] | 31.06 ± 12.27 | 193.89 [167.12] | 37.04 ± 7.12 |
| CDCA | 107.68 [283.84] | 9.37 ± 9.38 | 0.31 [0.10] | 0.07 ± 0.05 |
| gCDCA | 317.86 [506.23] | 19.74 ± 10.87 | 148.01 [145.83] | 27.56 ± 6.59 |
| tCDCA | 26.66 [48.57] | 1.95 ± 1.50 | 49.50 [39.29] | 9.41 ± 3.98 |
| Total DCA | 586.74 [738.24] | 31.99 ± 13.07 | 97.31 [138.25] | 19.43 ± 12.23 |
| DCA | 303.83 [365.36] | 17.51 ± 10.06 | 0.21 [0.09] | 0.04 ± 0.03 |
| gDCA | 152.38 [319.50] | 11.04 ± 7.14 | 74.38 [117.62] | 15.36 ± 10.22 |
| tDCA | 43.35 [76.15] | 3.44 ± 3.34 | 14.94 [30.31] | 4.03 ± 2.97 |
| Total HCA | 5.91 [6.96] | 0.45 ± 0.44 | 0.41 [0.34] | 0.09 ± 0.07 |
| HCA | 2.63 [5.09] | 0.25 ± 0.37 | ND | ND |
| gHCA | 2.57 [2.15] | 0.19 ± 0.25 | 0.22 [0.25] | 0.05 ± 0.04 |
| tHCA | ND | ND | 0.18 [0.09] | 0.04 ± 0.03 |
| Total HDCA | 218.96 [303.82] | 13.84 ± 9.79 | 0.05 [0.07] | 0.01 ± 0.01 |
| HDCA | 209.17 [301.29] | 13.65 ± 9.78 | ND | ND |
| gHDCA | 2.25 [4.38] | 0.19 ± 0.17 | 0.05 [0.07] | 0.01 ± 0.01 |
| Total UDCA | 195.23 [219.81] | 11.51 ± 6.88 | 13.38 [19.52] | 3.30 ± 3.29 |
| UDCA | 153.80 [168.33] | 8.99 ± 6.14 | 0.07 [0.06] | 0.02 ± 0.03 |
| gUDCA | 35.07 [49.80] | 2.41 ± 2.14 | 12.12 [17.71] | 2.94 ± 3.00 |
| tUDCA | 1.37 [2.34] | 0.11 ± 0.13 | 1.18 [1.40] | 0.34 ± 0.36 |
| Total DHCA | ND | ND | 0.14 [0.04] | 0.03 ± 0.02 |
| gDHCA | ND | ND | 0.02 [0.01] | 0.01 ± 0.00 |
| tDHCA | ND | ND | 0.11 [0.04] | 0.02 ± 0.01 |
| Total LCA | 22.02 [22.52] | 1.65 ± 1.58 | 6.07 [5.29] | 1.38 ± 1.04 |
| LCA | 8.06 [10.87] | 0.66 ± 0.74 | 0.94 [0.15] | 0.20 ± 0.11 |
| gLCA | 10.03 [10.82] | 0.82 ± 0.81 | 3.30 [4.15] | 0.86 ± 0.90 |
| tLCA | 2.14 [2.08] | 0.17 ± 0.14 | 1.27 [1.75] | 0.32 ± 0.27 |
| Total BA | 1845.60 [2073.32] | – | 516.23 [480.98] | – |
| Total primary | 763.34 [1133.66] | 41.02 ± 16.07 | 397.55 [306.38] | 75.76 ± 12.54 |
| Total secondary | 1198.78 [1149.99] | 58.98 ± 16.07 | 118.45 [145.36] | 24.15 ± 12.53 |
| Total unconjugated | 1034.55 [1131.51] | 54.09 ± 20.72 | 1.67 [0.37] | 0.36 ± 0.19 |
| Total conjugated | 753.82 [1178.53] | 45.91 ± 20.72 | 514.73 [480.73] | 99.64 ± 0.19 |
| Total G-conjugated | 662.00 [1003.90] | 40.24 ± 18.24 | 407.15 [372.23] | 76.02 ± 9.51 |
| Total T-conjugated | 80.98 [129.02] | 5.67 ± 4.57 | 119.32 [99.02] | 23.62 ± 9.48 |
| Abbreviations: CA, cholic acid; CDCA, chenodeoxycholic acid; DCA, deoxycholic acid; DHCA, dehydrocholic acid; HCA, hyocholic acid; HDCA, hyodeoxycholic acid; UDCA, ursodeoxycholic acid; LCA, lithocholic acid; ND, not detected. |  |  |  |  |
| BAs prefixed with 'g' denote glycine conjugates, and those with 't' denote taurine conjugates. |  |  |  |  |

**Table S3.** Clinical characteristics of participants with MASLD and controls in the main liver biopsy cohort.

|  | Controls | MASLD |
| --- | --- | --- |
| Group size, *n* | 26 | 77 |
| Age, years | 47 ± 2 | 49 ± 1 |
| Females, *n* (%) | 19 (73) | 50 (65) |
| BMI, kg/m<sup>2</sup> | 43.2 (40.2–49.3) | 45.4 (42.3–48.7) |
| Waist circumference, cm | 126 (115–128) | 134 (124–141)** |
| Systolic blood pressure, mmHg | 128 (118–144) | 135 (125–145) |
| Diastolic blood pressure, mmHg | 86 (83–95) | 92 (85–97) |
| Plasma glucose, mmol/L | 5.2 (4.7–5.7) | 6.0 (5.5–6.8)*** |
| HbA1c, % | 5.5 (5.4–5.7) | 6.0 (5.7–6.4)*** |
| Serum insulin, mIU/L | 6.6 (5.8–13.3) | 13.2 (8.4–18.4)*** |
| Serum C-peptide, nmol/L | 0.91 ± 0.09 | 1.14 ± 0.04* |
| HOMA-IR | 1.5 (1.2–3.2) | 3.4 (2.3–5.2)*** |
| Matsuda ISI<sup>†</sup> | 104.4 (53.3–170.3) | 47.3 (35.2–68.8)*** |
| Plasma total cholesterol, mmol/L | 4.4 (3.7–4.7) | 4.0 (3.4–4.8) |
| Plasma HDL cholesterol, mmol/L | 1.13 (1.02–1.46) | 1.08 (0.93–1.25) |
| Plasma LDL cholesterol, mmol/L | 2.6 (2.2–3.0) | 2.2 (1.6–2.9) |
| Plasma triglycerides, mmol/L | 0.97 (0.73–1.23) | 1.40 (1.11–1.91)*** |
| Plasma ALT, IU/L | 28 (23–36) | 35 (27–48)* |
| Plasma AST, IU/L | 26 (24–33) | 30 (26–37) |
| Plasma AST/ALT ratio | 0.95 (0.86–1.21) | 0.86 (0.73–1.08) |
| Plasma GGT, IU/L | 30 (17–43) | 31 (23–47) |
| Plasma ALP, IU/L | 64 ± 3 | 66 ± 2 |
| Plasma albumin, g/L | 38 ± 1 | 38 ± 1 |
| Platelet count, 10<sup>9</sup>/L | 233 (207–268) | 248 (209–299) |
| Serum adiponectin, µg/mL | 11.4 (8.7–13.8) | 6.4 (4.3–9.4)*** |
| IHTG content (histology), % | 0 (0–0) | 20 (10–30)*** |
| Fibrosis stage (F0/F1/F2), *n* | 23/1/2 | 40/34/3*** |
| Type 2 diabetes, *n* (%) | 5 (19) | 45 (58)*** |
| Use of metformin, *n* (%) | 4 (15) | 43 (56)*** |
| Use of statins, *n* (%) | 6 (23) | 31 (40) |
| Current smoking, *n* (%) | 6 (23) | 17 (22) |
| Genetic risk score<sup>‡</sup> | 0.19 ± 0.04 | 0.29 ± 0.02* |

Data are shown as *n* (%), mean ± SEM or median (25<sup>th</sup>–75<sup>th</sup> percentiles). Statistical tests used were the Student's *t*-test, Mann-Whitney *U* test, Pearson's $\chi^2$ test, or the Fisher's exact test, as appropriate. \**P* < 0.05; \*\**P* < 0.01; \*\*\**P* < 0.001.
<sup>†</sup>Available for *n* = 91.
<sup>‡</sup>Calculated using the PNPLA3 rs738409, TM6SF2 rs58542926, MBOAT7 rs641738, and GCKR rs1260326 genotypes as described in Methods.

**Table S4.** Concentrations of serum BAs in patients with MASLD and controls in the main liver biopsy cohort.

|  | Controls (*n* = 26) |  | MASLD (*n* = 77) |  |  |  |
| --- | --- | --- | --- | --- | --- | --- |
|  | Median | IQR | Median | IQR | *P* | *P*<sub>adj</sub> |
| Total CA | 81.33 | 125.74 | 128.97 | 217.24 | .018 | .043 |
| CA | 20.49 | 30.34 | 25.80 | 50.78 | .112 | .189 |
| gCA | 49.77 | 56.33 | 82.78 | 172.91 | .008 | .032 |
| tCA | ND | ND | ND | ND | – | – |
| Total CDCA | 358.07 | 392.20 | 663.97 | 851.94 | .004 | .032 |
| CDCA | 49.61 | 192.30 | 118.21 | 313.70 | .046 | .093 |
| gCDCA | 227.59 | 190.05 | 390.37 | 615.72 | .006 | .032 |
| tCDCA | 20.58 | 35.03 | 26.69 | 52.35 | .178 | .268 |
| Total DCA | 365.38 | 305.75 | 700.04 | 761.90 | .004 | .032 |
| DCA | 212.09 | 176.01 | 369.24 | 520.03 | .008 | .032 |
| gDCA | 93.24 | 203.23 | 198.36 | 336.44 | .037 | .079 |
| tDCA | 35.32 | 57.70 | 43.84 | 78.57 | .250 | .320 |
| Total HCA | 6.81 | 7.87 | 5.50 | 5.81 | .376 | .415 |
| HCA | 2.62 | 2.15 | 2.68 | 5.48 | .707 | .754 |
| gHCA | 2.91 | 2.56 | 2.29 | 2.29 | .097 | .173 |
| tHCA | ND | ND | ND | ND | – | – |
| Total HDCA | 173.55 | 211.65 | 236.78 | 337.18 | .019 | .043 |
| HDCA | 171.26 | 211.89 | 232.00 | 333.51 | .019 | .043 |
| gHDCA | 1.87 | 1.96 | 2.53 | 5.50 | .198 | .268 |
| Total UDCA | 177.61 | 248.19 | 199.23 | 210.12 | .311 | .383 |
| UDCA | 115.07 | 186.84 | 160.17 | 162.93 | .376 | .415 |
| gUDCA | 26.19 | 53.54 | 45.91 | 48.96 | .337 | .399 |
| tUDCA | 0.80 | 2.59 | 1.44 | 2.22 | .190 | .268 |
| Total DHCA | ND | ND | ND | ND | – | – |
| gDHCA | ND | ND | ND | ND | – | – |
| tDHCA | ND | ND | ND | ND | – | – |
| Total LCA | 19.79 | 10.77 | 22.93 | 25.59 | .201 | .268 |
| LCA | 6.22 | 6.96 | 8.99 | 13.94 | .079 | .148 |
| gLCA | 10.43 | 6.58 | 9.24 | 12.49 | .882 | .882 |
| tLCA | 2.15 | 2.06 | 2.10 | 2.27 | .764 | .789 |
| Total BA | 1326.09 | 1456.70 | 2340.33 | 2106.01 | .002 | .032 |
| Total primary | 415.61 | 487.71 | 812.35 | 1173.19 | .007 | .032 |
| Total secondary | 768.99 | 723.43 | 1297.72 | 1261.71 | .003 | .032 |
| Total unconjugated | 749.11 | 800.30 | 1139.89 | 1413.56 | .018 | .043 |
| Total conjugated | 496.25 | 630.62 | 945.26 | 1322.56 | .009 | .032 |
| Total G-conjugated | 430.04 | 552.26 | 835.28 | 1143.96 | .010 | .032 |
| Total T-conjugated | 66.51 | 115.75 | 83.01 | 123.16 | .173 | .268 |

Units are nmol/L. The Mann-Whitney *U* test was used to test for differences between the groups, and false discovery rate was controlled with Benjamini-Hochberg-adjusted *P*-values.
Abbreviations: CA, cholic acid; CDCA, chenodeoxycholic acid; DCA, deoxycholic acid; DHCA, dehydrocholic acid; HCA, hyocholic acid; HDCA, hyodeoxycholic acid; UDCA, ursodeoxycholic acid; LCA, lithocholic acid; ND, not detected.
BAs prefixed with 'g' denote glycine conjugates, and those with 't' denote taurine conjugates.

**Table S5.** Serum BA concentrations in groups based on sex-specific medians of HOMA-IR and the GRS in the main liver biopsy cohort.

|  |  |  |  |  |  |  |  |  |  |  |  |  |
| --- | --- | --- | --- | --- | --- | --- | --- | --- | --- | --- | --- | --- |
|  | Low HOMA-IR |  | High HOMA-IR |  |  |  | Low GRS |  | High GRS |  |  |  |
|  | Median | IQR | Median | IQR | *P* | *P*<sub>adj</sub> | Median | IQR | Median | IQR | *P* | *P*<sub>adj</sub> |
| Total CA | 91.94 | 131.03 | 132.73 | 223.70 | .013 | .037 | 124.42 | 229.97 | 94.80 | 168.76 | .290 | .987 |
| CA | 22.14 | 37.78 | 25.05 | 47.64 | .195 | .315 | 26.97 | 50.49 | 22.14 | 37.42 | .139 | .987 |
| gCA | 67.08 | 62.48 | 105.79 | 220.16 | .002 | .016 | 83.85 | 106.22 | 67.81 | 144.54 | .378 | .987 |
| Total CDCA | 398.11 | 687.25 | 718.92 | 1038.81 | .007 | .026 | 581.60 | 842.79 | 567.41 | 818.85 | .987 | .987 |
| CDCA | 71.74 | 257.85 | 117.40 | 285.52 | .140 | .263 | 98.64 | 412.76 | 116.59 | 246.83 | .600 | .987 |
| gCDCA | 266.88 | 334.15 | 413.86 | 676.31 | .011 | .034 | 326.12 | 484.57 | 303.56 | 482.31 | .877 | .987 |
| tCDCA | 18.71 | 20.53 | 44.87 | 53.88 | <.001 | <.001 | 26.40 | 44.51 | 26.66 | 46.97 | .764 | .987 |
| Total DCA | 483.20 | 557.97 | 838.46 | 823.46 | .022 | .051 | 568.03 | 727.65 | 600.60 | 731.30 | .892 | .987 |
| DCA | 283.96 | 281.92 | 366.32 | 510.70 | .275 | .400 | 301.94 | 468.21 | 308.97 | 363.26 | .929 | .987 |
| gDCA | 130.01 | 236.11 | 282.35 | 480.04 | .017 | .044 | 155.31 | 293.62 | 144.40 | 343.71 | .714 | .987 |
| tDCA | 27.48 | 44.52 | 64.45 | 102.62 | <.001 | .002 | 42.63 | 75.72 | 43.84 | 72.54 | .919 | .987 |
| Total HCA | 5.69 | 7.26 | 6.50 | 5.84 | .546 | .647 | 6.50 | 7.01 | 5.37 | 5.82 | .396 | .987 |
| HCA | 2.61 | 4.72 | 2.71 | 4.80 | .442 | .544 | 2.63 | 3.98 | 2.68 | 5.44 | .955 | .987 |
| gHCA | 2.39 | 2.36 | 2.65 | 2.22 | .368 | .471 | 2.81 | 2.13 | 2.19 | 2.50 | .107 | .987 |
| Total HDCA | 219.22 | 309.72 | 210.48 | 288.85 | .992 | .992 | 186.58 | 307.53 | 239.38 | 264.11 | .550 | .987 |
| HDCA | 215.51 | 309.50 | 200.63 | 287.23 | .961 | .992 | 184.27 | 303.35 | 235.99 | 265.14 | .546 | .987 |
| gHDCA | 2.53 | 3.14 | 1.97 | 5.61 | .749 | .799 | 2.26 | 5.03 | 1.96 | 3.47 | .987 | .987 |
| Total UDCA | 194.04 | 227.28 | 199.89 | 202.90 | .368 | .471 | 190.18 | 251.41 | 195.23 | 198.26 | .784 | .987 |
| UDCA | 126.04 | 224.60 | 154.05 | 143.63 | .614 | .702 | 131.32 | 213.03 | 160.87 | 145.53 | .623 | .987 |
| gUDCA | 28.23 | 53.69 | 46.55 | 52.38 | .148 | .263 | 43.60 | 56.27 | 25.45 | 44.22 | .122 | .987 |
| tUDCA | 0.96 | 1.29 | 1.84 | 3.91 | .003 | .016 | 1.26 | 2.52 | 1.42 | 1.76 | .950 | .987 |
| Total LCA | 19.69 | 19.96 | 24.88 | 21.92 | .197 | .315 | 19.92 | 24.62 | 24.13 | 21.83 | .799 | .987 |
| LCA | 7.28 | 7.28 | 12.14 | 13.03 | .258 | .393 | 8.02 | 9.31 | 8.29 | 11.58 | .955 | .987 |
| gLCA | 10.40 | 7.58 | 9.23 | 12.66 | .724 | .799 | 9.08 | 14.01 | 10.74 | 10.21 | .866 | .987 |
| tLCA | 1.88 | 1.59 | 2.27 | 2.53 | .066 | .141 | 2.15 | 1.67 | 2.00 | 2.78 | .934 | .987 |
| Total BA | 1660.93 | 1951.29 | 2472.16 | 2096.58 | .018 | .044 | 1821.22 | 2355.04 | 2041.23 | 1912.14 | .892 | .987 |
| Total primary | 542.61 | 782.13 | 895.63 | 1322.61 | .007 | .026 | 801.87 | 1166.18 | 681.18 | 1078.53 | .774 | .987 |
| Total secondary | 940.49 | 1020.20 | 1337.17 | 1268.95 | .079 | .158 | 1114.99 | 1088.97 | 1255.36 | 1105.64 | .495 | .987 |
| Total unconjugated | 872.62 | 1017.93 | 1127.57 | 1222.93 | .341 | .471 | 923.94 | 1357.36 | 1067.24 | 982.73 | .810 | .987 |
| Total conjugated | 624.72 | 650.92 | 1122.85 | 1625.68 | .002 | .016 | 748.72 | 1192.31 | 753.82 | 1125.03 | .734 | .987 |
| Total G-conjugated | 590.81 | 591.16 | 1010.57 | 1526.37 | .006 | .026 | 705.71 | 901.40 | 657.81 | 1047.54 | .675 | .987 |
| Total T-conjugated | 45.64 | 61.71 | 130.09 | 150.39 | <.001 | <.001 | 73.45 | 119.20 | 83.01 | 129.46 | .835 | .987 |

Units are nmol/L. The Mann-Whitney *U* test was used to test for differences between the groups, and false discovery rate was controlled with Benjamini-Hochberg-adjusted *P*-values.
Abbreviations: CA, cholic acid; CDCA, chenodeoxycholic acid; DCA, deoxycholic acid; DHCA, dehydrocholic acid; HCA, hyocholic acid; HDCA, hyodeoxycholic acid; UDCA, ursodeoxycholic acid; LCA, lithocholic acid.
BAs prefixed with 'g' denote glycine conjugates, and those with 't' denote taurine conjugates.

**Table S6.**
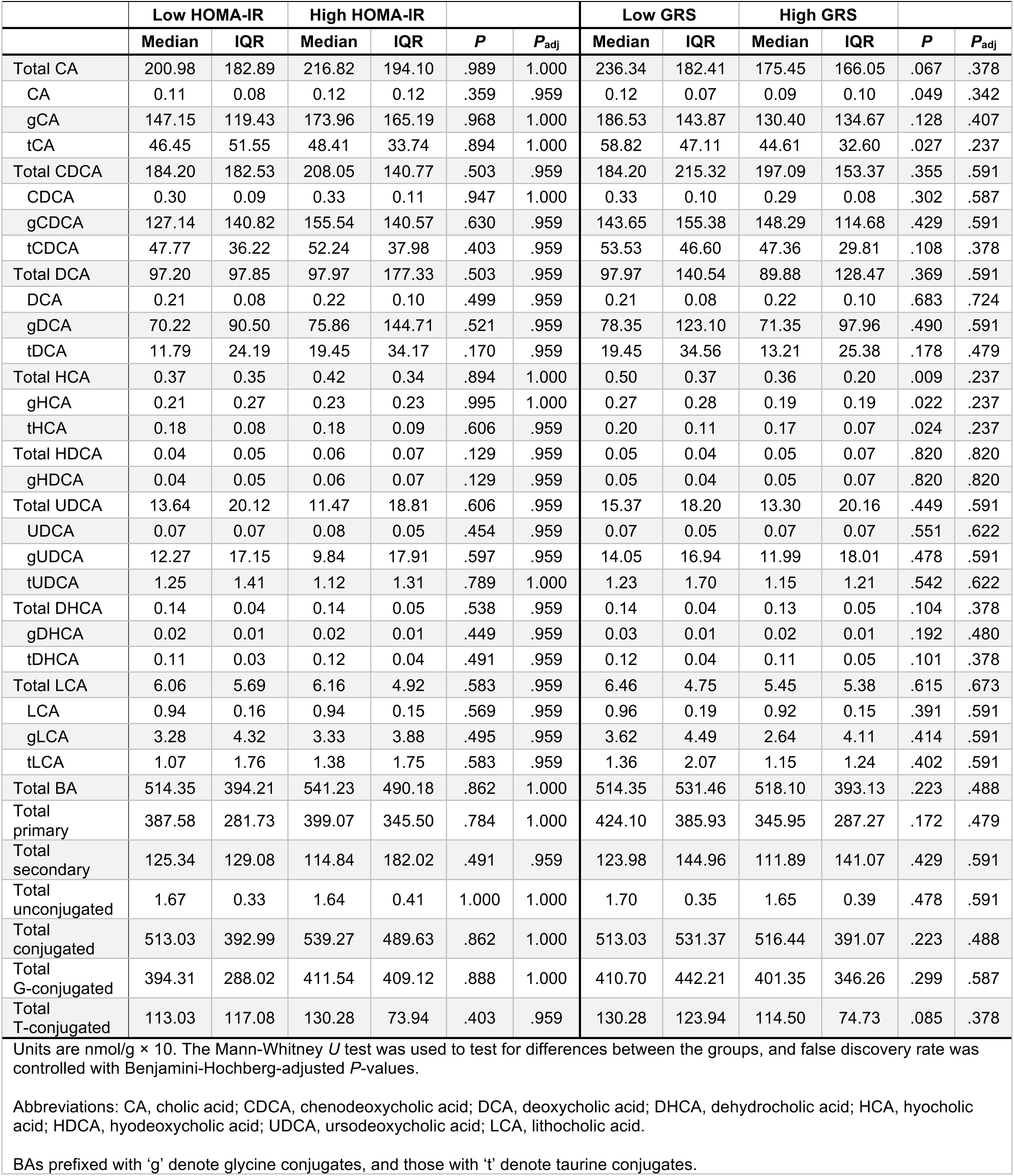
Liver BA concentrations in groups based on sex-specific medians of HOMA-IR and the GRS in the main liver biopsy cohort.

|  | Low HOMA-IR |  | High HOMA-IR |  |  |  | Low GRS |  | High GRS |  |  |  |
| --- | --- | --- | --- | --- | --- | --- | --- | --- | --- | --- | --- | --- |
|  | Median | IQR | Median | IQR | P | P<sub>adj</sub> | Median | IQR | Median | IQR | P | P<sub>adj</sub> |
| Total CA | 200.98 | 182.89 | 216.82 | 194.10 | .989 | 1.000 | 236.34 | 182.41 | 175.45 | 166.05 | .067 | .378 |
| CA | 0.11 | 0.08 | 0.12 | 0.12 | .359 | .959 | 0.12 | 0.07 | 0.09 | 0.10 | .049 | .342 |
| gCA | 147.15 | 119.43 | 173.96 | 165.19 | .968 | 1.000 | 186.53 | 143.87 | 130.40 | 134.67 | .128 | .407 |
| tCA | 46.45 | 51.55 | 48.41 | 33.74 | .894 | 1.000 | 58.82 | 47.11 | 44.61 | 32.60 | .027 | .237 |
| Total CDCA | 184.20 | 182.53 | 208.05 | 140.77 | .503 | .959 | 184.20 | 215.32 | 197.09 | 153.37 | .355 | .591 |
| CDCA | 0.30 | 0.09 | 0.33 | 0.11 | .947 | 1.000 | 0.33 | 0.10 | 0.29 | 0.08 | .302 | .587 |
| gCDCA | 127.14 | 140.82 | 155.54 | 140.57 | .630 | .959 | 143.65 | 155.38 | 148.29 | 114.68 | .429 | .591 |
| tCDCA | 47.77 | 36.22 | 52.24 | 37.98 | .403 | .959 | 53.53 | 46.60 | 47.36 | 29.81 | .108 | .378 |
| Total DCA | 97.20 | 97.85 | 97.97 | 177.33 | .503 | .959 | 97.97 | 140.54 | 89.88 | 128.47 | .369 | .591 |
| DCA | 0.21 | 0.08 | 0.22 | 0.10 | .499 | .959 | 0.21 | 0.08 | 0.22 | 0.10 | .683 | .724 |
| gDCA | 70.22 | 90.50 | 75.86 | 144.71 | .521 | .959 | 78.35 | 123.10 | 71.35 | 97.96 | .490 | .591 |
| tDCA | 11.79 | 24.19 | 19.45 | 34.17 | .170 | .959 | 19.45 | 34.56 | 13.21 | 25.38 | .178 | .479 |
| Total HCA | 0.37 | 0.35 | 0.42 | 0.34 | .894 | 1.000 | 0.50 | 0.37 | 0.36 | 0.20 | .009 | .237 |
| gHCA | 0.21 | 0.27 | 0.23 | 0.23 | .995 | 1.000 | 0.27 | 0.28 | 0.19 | 0.19 | .022 | .237 |
| tHCA | 0.18 | 0.08 | 0.18 | 0.09 | .606 | .959 | 0.20 | 0.11 | 0.17 | 0.07 | .024 | .237 |
| Total HDCA | 0.04 | 0.05 | 0.06 | 0.07 | .129 | .959 | 0.05 | 0.04 | 0.05 | 0.07 | .820 | .820 |
| gHDCA | 0.04 | 0.05 | 0.06 | 0.07 | .129 | .959 | 0.05 | 0.04 | 0.05 | 0.07 | .820 | .820 |
| Total UDCA | 13.64 | 20.12 | 11.47 | 18.81 | .606 | .959 | 15.37 | 18.20 | 13.30 | 20.16 | .449 | .591 |
| UDCA | 0.07 | 0.07 | 0.08 | 0.05 | .454 | .959 | 0.07 | 0.05 | 0.07 | 0.07 | .551 | .622 |
| gUDCA | 12.27 | 17.15 | 9.84 | 17.91 | .597 | .959 | 14.05 | 16.94 | 11.99 | 18.01 | .478 | .591 |
| tUDCA | 1.25 | 1.41 | 1.12 | 1.31 | .789 | 1.000 | 1.23 | 1.70 | 1.15 | 1.21 | .542 | .622 |
| Total DHCA | 0.14 | 0.04 | 0.14 | 0.05 | .538 | .959 | 0.14 | 0.04 | 0.13 | 0.05 | .104 | .378 |
| gDHCA | 0.02 | 0.01 | 0.02 | 0.01 | .449 | .959 | 0.03 | 0.01 | 0.02 | 0.01 | .192 | .480 |
| tDHCA | 0.11 | 0.03 | 0.12 | 0.04 | .491 | .959 | 0.12 | 0.04 | 0.11 | 0.05 | .101 | .378 |
| Total LCA | 6.06 | 5.69 | 6.16 | 4.92 | .583 | .959 | 6.46 | 4.75 | 5.45 | 5.38 | .615 | .673 |
| LCA | 0.94 | 0.16 | 0.94 | 0.15 | .569 | .959 | 0.96 | 0.19 | 0.92 | 0.15 | .391 | .591 |
| gLCA | 3.28 | 4.32 | 3.33 | 3.88 | .495 | .959 | 3.62 | 4.49 | 2.64 | 4.11 | .414 | .591 |
| tLCA | 1.07 | 1.76 | 1.38 | 1.75 | .583 | .959 | 1.36 | 2.07 | 1.15 | 1.24 | .402 | .591 |
| Total BA | 514.35 | 394.21 | 541.23 | 490.18 | .862 | 1.000 | 514.35 | 531.46 | 518.10 | 393.13 | .223 | .488 |
| Total primary | 387.58 | 281.73 | 399.07 | 345.50 | .784 | 1.000 | 424.10 | 385.93 | 345.95 | 287.27 | .172 | .479 |
| Total secondary | 125.34 | 129.08 | 114.84 | 182.02 | .491 | .959 | 123.98 | 144.96 | 111.89 | 141.07 | .429 | .591 |
| Total unconjugated | 1.67 | 0.33 | 1.64 | 0.41 | 1.000 | 1.000 | 1.70 | 0.35 | 1.65 | 0.39 | .478 | .591 |
| Total conjugated | 513.03 | 392.99 | 539.27 | 489.63 | .862 | 1.000 | 513.03 | 531.37 | 516.44 | 391.07 | .223 | .488 |
| Total G-conjugated | 394.31 | 288.02 | 411.54 | 409.12 | .888 | 1.000 | 410.70 | 442.21 | 401.35 | 346.26 | .299 | .587 |
| Total T-conjugated | 113.03 | 117.08 | 130.28 | 73.94 | .403 | .959 | 130.28 | 123.94 | 114.50 | 74.73 | .085 | .378 |

Units are nmol/g $\times 10$ . The Mann-Whitney *U* test was used to test for differences between the groups, and false discovery rate was controlled with Benjamini-Hochberg-adjusted *P*-values.
Abbreviations: CA, cholic acid; CDCA, chenodeoxycholic acid; DCA, deoxycholic acid; DHCA, dehydrocholic acid; HCA, hyocholic acid; HDCA, hyodeoxycholic acid; UDCA, ursodeoxycholic acid; LCA, lithocholic acid.
BAs prefixed with 'g' denote glycine conjugates, and those with 't' denote taurine conjugates.

**Table S7.** Multiple linear regression analysis of serum BA concentrations, HOMA-IR, and GRS in the main liver biopsy cohort.

|  |  |  |  |  |  |  |  |  |
| --- | --- | --- | --- | --- | --- | --- | --- | --- |
|  | **HOMA-IR** |  |  |  | **GRS** |  |  |  |
| | $\beta$ | SE | 95% CI | *P* | $\beta$ | SE | 95% CI | *P* |
| Total CA | 0.582 | 0.191 | 0.203–0.960 | .003 | -0.049 | 0.052 | -0.153–0.055 | .351 |
| CA | 0.603 | 0.243 | 0.120–1.087 | .015 | -0.041 | 0.067 | -0.173–0.092 | .543 |
| gCA | 0.457 | 0.183 | 0.093–0.820 | .014 | -0.048 | 0.050 | -0.147–0.052 | .345 |
| Total CDCA | 0.386 | 0.163 | 0.063–0.709 | .020 | -0.026 | 0.045 | -0.115–0.063 | .559 |
| CDCA | 0.442 | 0.260 | -0.074–0.958 | .092 | 0.007 | 0.071 | -0.135–0.149 | .922 |
| gCDCA | 0.353 | 0.167 | 0.022–0.684 | .037 | -0.049 | 0.046 | -0.140–0.042 | .287 |
| tCDCA | 0.579 | 0.177 | 0.227–0.931 | .002 | -0.038 | 0.049 | -0.134–0.059 | .438 |
| Total DCA | 0.315 | 0.161 | -0.005–0.636 | .053 | 0.001 | 0.044 | -0.087–0.089 | .987 |
| DCA | 0.197 | 0.199 | -0.198–0.592 | .325 | -0.003 | 0.055 | -0.112–0.105 | .952 |
| gDCA | 0.390 | 0.208 | -0.023–0.804 | .064 | -0.018 | 0.057 | -0.132–0.095 | .749 |
| tDCA | 0.526 | 0.199 | 0.131–0.922 | .010 | -0.064 | 0.055 | -0.172–0.045 | .246 |
| Total HCA | 0.128 | 0.166 | -0.203–0.458 | .444 | 0.010 | 0.046 | -0.080–0.101 | .822 |
| HCA | 0.153 | 0.226 | -0.296–0.602 | .500 | 0.027 | 0.062 | -0.096–0.151 | .659 |
| gHCA | 0.186 | 0.160 | -0.132–0.504 | .249 | -0.064 | 0.044 | -0.151–0.023 | .148 |
| Total HDCA | -0.086 | 0.153 | -0.390–0.219 | .577 | 0.026 | 0.042 | -0.058–0.109 | .542 |
| HDCA | -0.085 | 0.154 | -0.392–0.221 | .582 | 0.026 | 0.042 | -0.059–0.110 | .549 |
| gHDCA | -0.252 | 0.224 | -0.696–0.192 | .263 | 0.024 | 0.061 | -0.098–0.146 | .694 |
| Total UDCA | 0.091 | 0.164 | -0.236–0.417 | .582 | 0.038 | 0.045 | -0.052–0.127 | .406 |
| UDCA | 0.022 | 0.178 | -0.332–0.376 | .904 | 0.047 | 0.049 | -0.050–0.144 | .340 |
| gUDCA | 0.257 | 0.220 | -0.181–0.695 | .246 | -0.029 | 0.060 | -0.149–0.091 | .628 |
| tUDCA | 0.454 | 0.199 | 0.059–0.848 | .025 | -0.023 | 0.055 | -0.131–0.085 | .672 |
| Total LCA | 0.106 | 0.134 | -0.159–0.371 | .430 | -0.022 | 0.037 | -0.095–0.050 | .544 |
| LCA | 0.152 | 0.170 | -0.185–0.489 | .374 | -0.022 | 0.047 | -0.114–0.071 | .640 |
| gLCA | 0.076 | 0.144 | -0.210–0.362 | .599 | -0.022 | 0.040 | -0.100–0.057 | .584 |
| tLCA | 0.104 | 0.122 | -0.138–0.346 | .394 | -0.046 | 0.033 | -0.112–0.020 | .172 |
| Total BA | 0.290 | 0.134 | 0.024–0.555 | .033 | -0.006 | 0.037 | -0.079–0.067 | .869 |
| Total primary | 0.434 | 0.164 | 0.108–0.760 | .010 | -0.029 | 0.045 | -0.118–0.060 | .520 |
| Total secondary | 0.202 | 0.132 | -0.060–0.465 | .129 | 0.016 | 0.036 | -0.056–0.088 | .662 |
| Total unconjugated | 0.181 | 0.157 | -0.131–0.492 | .253 | 0.009 | 0.043 | -0.077–0.095 | .834 |
| Total conjugated | 0.386 | 0.160 | 0.067–0.704 | .018 | -0.043 | 0.044 | -0.130–0.045 | .334 |
| Total G-conjugated | 0.356 | 0.163 | 0.033–0.679 | .031 | -0.043 | 0.045 | -0.132–0.045 | .333 |
| Total T-conjugated | 0.532 | 0.171 | 0.193–0.871 | .002 | -0.047 | 0.047 | -0.140–0.046 | .319 |

Linear models were adjusted for age, sex, BMI, type 2 diabetes, smoking status, and use of anti-diabetic and lipid-lowering medications. HOMA-IR was adjusted for the GRS and vice versa. The dependent variables (*i.e.*, BA concentrations) and independent variables (*i.e.*, HOMA-IR and GRS) underwent log-transformation prior to model building. Beta coefficients may be interpreted as % change in BA concentrations per 1% increase in HOMA-IR or GRS.
Abbreviations: CA, cholic acid; CDCA, chenodeoxycholic acid; DCA, deoxycholic acid; DHCA, dehydrocholic acid; HCA, hyocholic acid; HDCA, hyodeoxycholic acid; UDCA, ursodeoxycholic acid; LCA, lithocholic acid.
BAs prefixed with 'g' denote glycine conjugates, and those with 't' denote taurine conjugates.

**Table S8.**
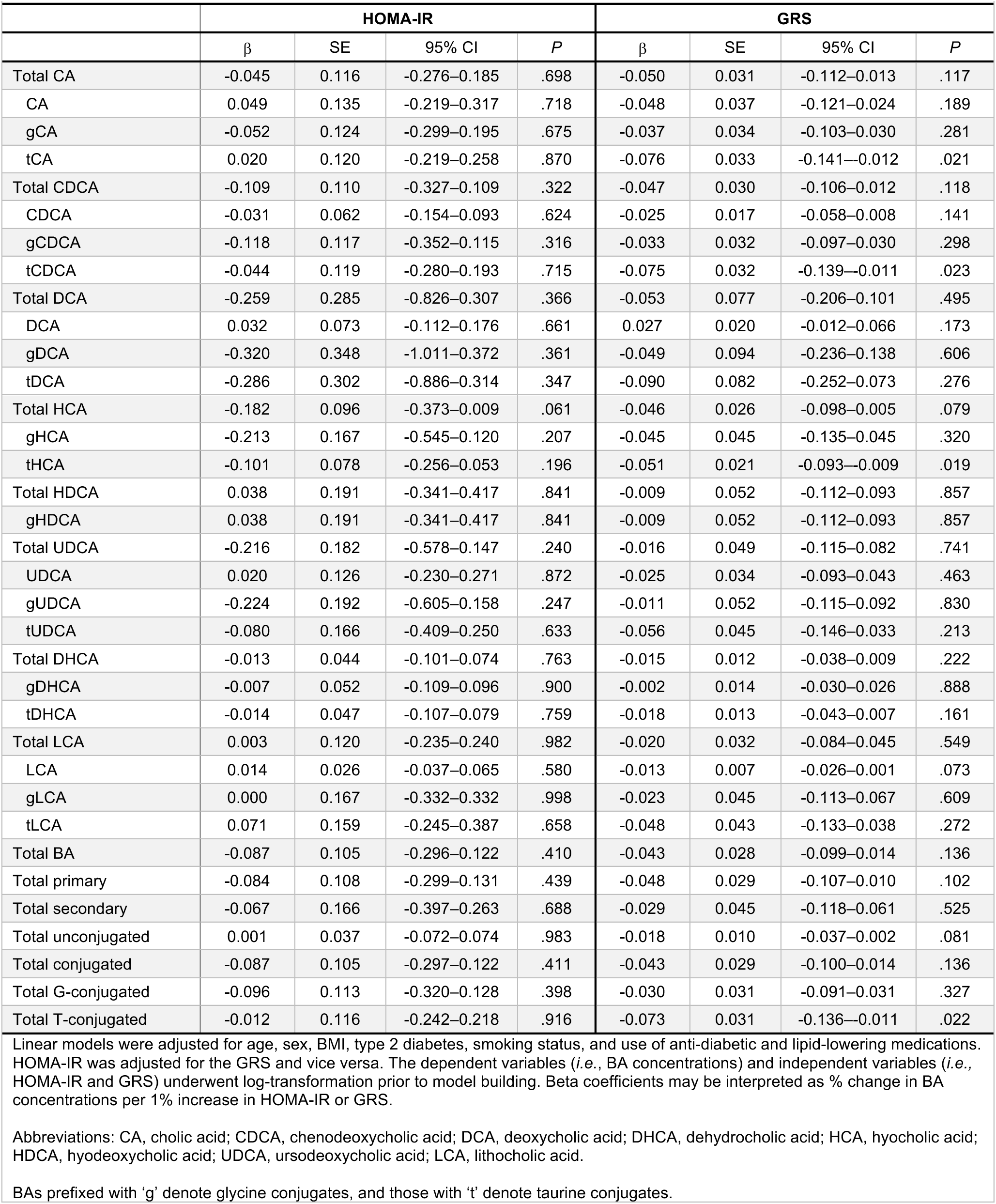
Multiple linear regression analysis of liver BA concentrations, HOMA-IR, and GRS in the main liver biopsy cohort.

**Table S9.**
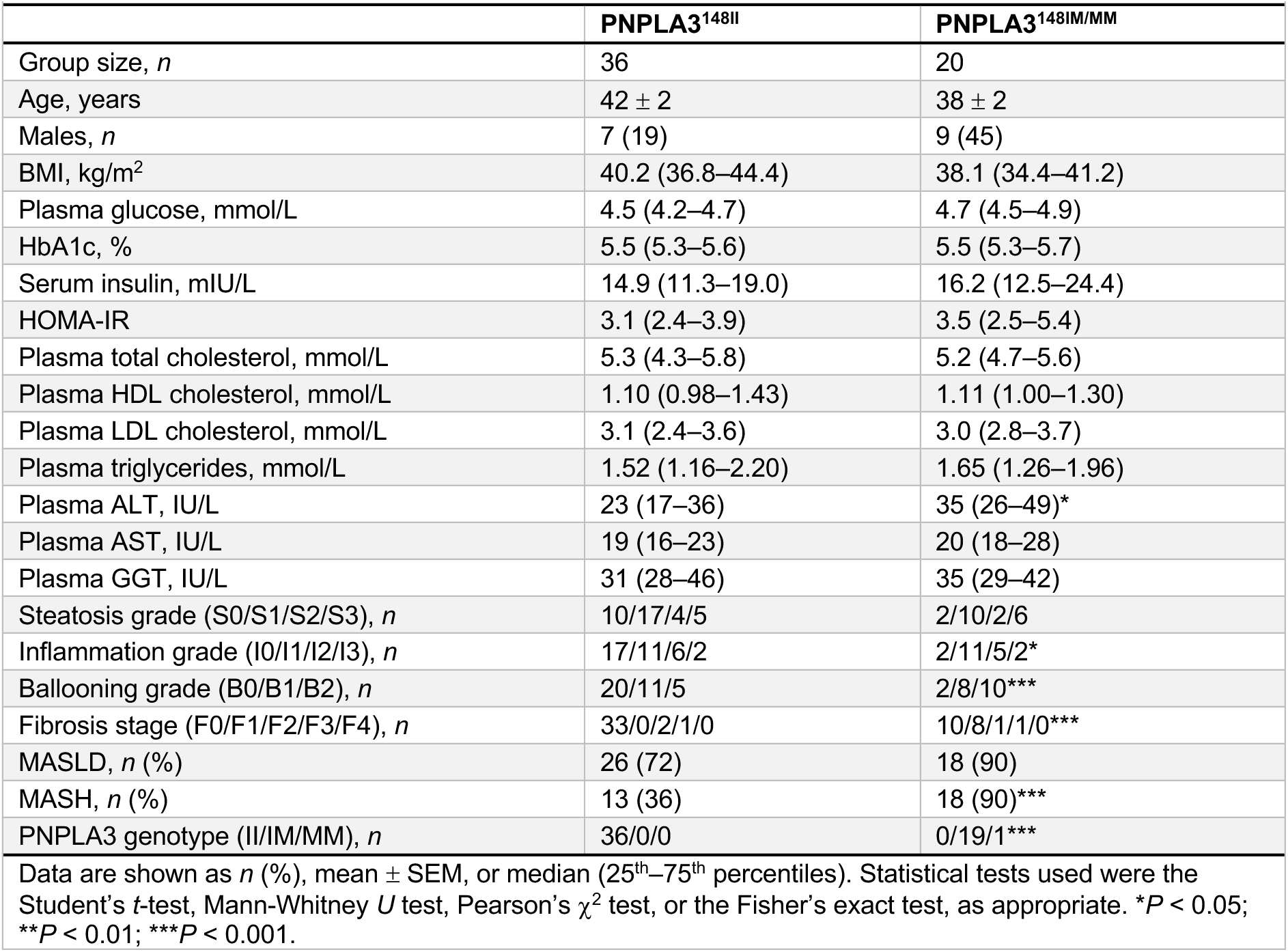
Clinical characteristics of PNPLA3-I148M carriers and non-carriers in the Antwerp validation cohort.

**Table S10.**
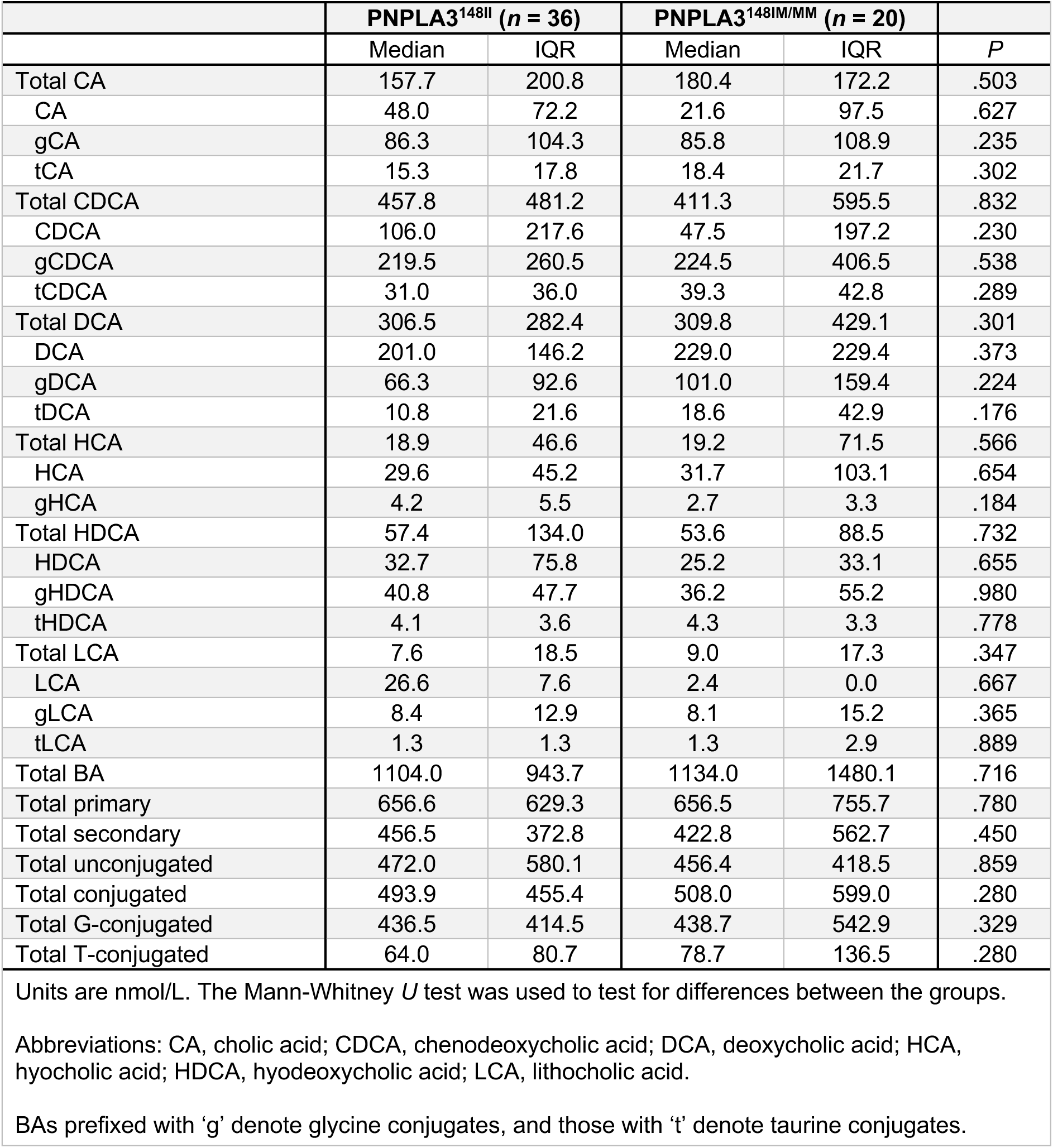
Concentrations of serum BAs in PNPLA3-I148M carriers and non-carriers in the Antwerp validation cohort.

**Table S11.** Clinical characteristics of the two liver RNA-seq cohorts.

|  | Derivation cohort | Validation cohort |
| --- | --- | --- |
| Group size, *n* | 127 | 86 |
| Age, years | 50 ± 1 | 47 ± 1 |
| Females, *n* (%) | 87 (69) | 60 (70) |
| BMI, kg/m<sup>2</sup> | 42.4 (37.7–46.8) | 45.6 (41.2–49.1) |
| Waist circumference, cm | 126 (112–135) | 130 (120–138) |
| Systolic blood pressure, mmHg | 128 (118–137) | 132 (122–143) |
| Diastolic blood pressure, mmHg | 84 (77–93) | 92 (85–95) |
| Plasma glucose, mmol/L | 5.9 (5.3–6.5) | 5.7 (5.1–6.2) |
| HbA1c, % | 5.8 (5.4–6.3) | 5.7 (5.5–6.2) |
| Serum insulin, mIU/L | 11.1 (7.1–16.7) | 11.8 (7.4–18.8) |
| Serum C-peptide, nmol/L | 0.94 ± 0.04 | 1.08 ± 0.05 |
| HOMA-IR | 3.0 (1.9–4.6) | 3.2 (1.7–5.1) |
| Plasma total cholesterol, mmol/L | 3.9 (3.6–4.5) | 4.2 (3.7–4.8) |
| Plasma HDL cholesterol, mmol/L | 1.12 (0.98–1.35) | 1.12 (0.96–1.36) |
| Plasma LDL cholesterol, mmol/L | 2.4 (1.9–2.7) | 2.4 (1.9–3.1) |
| Plasma triglycerides, mmol/L | 1.20 (0.92–1.71) | 1.23 (0.94–1.69) |
| Plasma ALT, IU/L | 31 (22–45) | 33 (24–45) |
| Plasma AST, IU/L | 28 (23–35) | 31 (25–38) |
| Plasma AST/ALT ratio | 0.92 (0.73–1.10) | 0.91 (0.74–1.18) |
| Plasma GGT, IU/L | 26 (19–46) | 29 (20–46) |
| Plasma ALP, IU/L | 65 ± 2 | 66 ± 2 |
| Plasma albumin, g/L | 38 ± 3 | 38 ± 1 |
| Platelet count, 10<sup>9</sup>/L | 251 (220–283) | 254 (218–292) |
| IHTG content (histology), % | 5 (0–20) | 5 (0–25) |
| Fibrosis stage (F0/F1/F2), *n* | 73/48/6 | 55/28/3 |
| MASLD, *n* (%) | 82 (65) | 58 (67) |
| MASH, *n* (%) | 16 (13) | 7 (8) |
| Type 2 diabetes, *n* (%) | 75 (59) | 27 (31) |
| Use of metformin, *n* (%) | 74 (58) | 25 (29) |
| Use of statins, *n* (%) | 44 (35) | 26 (30) |
| Current smoking, *n* (%) | 14 (11) | 20 (23) |
| Genetic risk score<sup>†</sup> | 0.27 ± 0.02 | 0.27 ± 0.02 |

Data are shown as *n* (%), mean ± SEM, or median (25<sup>th</sup>–75<sup>th</sup> percentiles).
<sup>†</sup>Calculated using the PNPLA3 rs738409, TM6SF2 rs58542926, MBOAT7 rs641738, and GCKR rs1260326 genotypes as described in Methods.

**Table S12.** A list of key genes related to different aspects of hepatic BA metabolism.

| Bile acid biosynthesis | Signaling regulation | Conjugation and detoxification | Bile secretion |  | Transport |
| --- | --- | --- | --- | --- | --- |
| CYP7A1 | NR1H4 | BAAT | ABCB1 | UGT1A3 | SLC10A1 |
| CYP27A1 | NR0B2 | SLC27A2 | ABCB4 | UGT2B10 | EPHX1 |
| CYP7B1 | RXRA | SLC27A5 | AQP1 | UGT1A9 | SLCO1C1 |
| CH25H | NR5A2 | ACOT8 | AQP4 | UGT1A10 | SLCO4A1 |
| CYP8B1 | FGF19 | CYP3A4 | AQP8 | UGT1A8 | SLCO1A2 |
| CYP39A1 | MALRD1 | CYP2B6 | AQP9 | UGT1A5 | SLCO1B3 |
| CYP46A1 | KNG1 | CYP2C9 | GNAS | UGT2B15 | SLCO2B1 |
| AKR1D1 | FGFR4 | SULT2A1 | SCTR | UGT1A7 | SLCO3A1 |
| AKR1C4 | KLB | UGT1A1 | SLC4A2 | UGT2A2 | SLCO4C1 |
| HSD3B7 | HNF1A | UGT2B4 | SLC4A4 | SLC22A7 | SLCO6A1 |
| ACOX2 | HNF4A | UGT2B7 | SLC10A2 | SLC22A8 | FABP6 |
| HSD17B4 | NR1I2 |  | SLC4A5 | SCARB1 | ABCB11 |
| SCP2 | VDR |  | KCNN2 | LDLR | ABCC2 |
| AMACR | GPBAR1 |  | ABCG1 | NCEH1 | ABCC3 |
|  | PPARA |  | ABCG2 | SLC9A1 | ABCC4 |
|  | RARA |  | CA2 | SCT | ABCC5 |
|  | NR1H3 |  | ATP8B1 | ADCY1 | SLC51A |
|  | RXRB |  | SLC5A1 | ADCY2 | SLC51B |
|  | NR1H2 |  | SLC22A1 | ADCY3 | SLCO1B1 |
|  | RXRG |  | SLC9A2 | ADCY4 | ABCC1 |
|  |  |  | SLC9A3 | ADCY5 |  |
|  |  |  | ABCG5 | ADCY6 |  |
|  |  |  | ABCG8 | ADCY7 |  |
|  |  |  | UGT2A1 | ADCY8 |  |
|  |  |  | UGT2A3 | ADCY9 |  |
|  |  |  | UGT2B17 | PRKACA |  |
|  |  |  | UGT2B11 | CFTR |  |
|  |  |  | UGT2B28 | SLC2A1 |  |
|  |  |  | UGT1A6 | PRKACB |  |
|  |  |  | UGT1A4 | PRKACG |  |

Gene lists were compiled based on the literature and the Kyoto Encyclopaedia of Genes and Genomes (KEGG) pathway database.

